# Application of an Age-Structured Deterministic Endemic Model for Disease Control in Nigeria

**DOI:** 10.1101/2020.03.28.20046300

**Authors:** A. O. Victor, H. K. Oduwole

## Abstract

This paper focuses on the development and analysis of the endemic model for disease control in an aged-structured population in Nigeria. Upon the model framework development, the model equations were transformed into proportions with rate of change of the different compartments forming the model, thereby reducing the model equations from twelve to ten homogenous ordinary differential equations. The model exhibits two equilibria, the endemic state and the disease-free equilibrium state while successfully achieving a Reproductive Number *R*_0_ = 0.

The deterministic endemic susceptible-exposed-infected-removed-undetectable=untransmissible-susceptible (SEIRUS) model is analyzed for the existence and stability of the disease-free equilibrium state. We established that a disease-free equilibrium state exists and is locally asymptotically stable when the basic reproduction number 0 ≤ *R*_0_ < 1. Furthermore, numerical simulations were carried to complement the analytical results in investigating the effect treatment rate and the net transmission rate on recovery for both juvenile and adult sub-population in an age-structured population.

## 1. INTRODUCTION

The estimation of the rate of prevalence of HIV/AIDS in Nigeria is very key for the administration of relevant interventions by relevant stakeholders in the health sector and the Nigerian Center for Disease Control (NCDC) hence in this paper we develop and evaluate the new deterministic endemic age-structured SEIRUS compartmental model of the HIV/AIDS dynamics in Nigeria and a two-age-structured population framework for a deterministic endemic model is constructed for the development of an endemic deterministic model with Undetectable=Untrasmitable viral load compartment.

In recent study by [1] and an earliest study by [2] among others have been very motivational for this study and were focused on the epidemiology of HIV/AIDS using various models like SIR, SEIR, SIRS, SICA models formulated for epidemic case of disease control. However, because these earliest models did not take into account the endemicity of the diseases, this study therefore focuses on the endemic nature, developing and evaluating a deterministic endemic model for the control of further spread and eventual eradication of the disease in Nigeria.

## 2. THE MODEL VARIABLES AND PARAMETERS

The model variables and parameters for the investigation of the stability analysis of the equilibrium state for the new deterministic endemic model which is a motivation from [1] is given by;

| Variable | Description |
| --- | --- |
| $S_1(t)$ | Number of susceptible juveniles at time $t$ |
| $S_2(t)$ | Number of susceptible adult at time $t$ |
| $E_1(t)$ | Number of exposed juvenile at time $t$ |
| $E_2(t)$ | Number of exposed adults at time $t$ |
| $I_1(t)$ | Number of infected juveniles at time $t$ |
| $I_2(t)$ | Number of infected adults at time $t$ |
| $R_1(t)$ | Number of infected juveniles receiving HAART at time $t$ |
| $R_2(t)$ | Number of infected adults receiving HAART at time $t$ |
| $U_1(t)$ | Number of recovered juveniles satisfying U=U case at time $t$ |
| $U_2(t)$ | Number of recovered adults satisfying U=U case at time $t$ |
| $A_1(t)$ | Number of AIDS cases in the juvenile sub-population at time $t$ |
| $A_2(t)$ | Number of AIDS cases in the adult sub-population at time $t$ |
| Parameter | Description |
| $\nu_1$ | The rate at which HIV infected juveniles becomes AIDS patient. |
| $\nu_2$ | The rate at which HIV infected adults becomes AIDS patient. |
| $\lambda$ | Birth rate of the adult sub-population |
| $\mu_1$ | Natural death rate of the juvenile sub-population |
| $\mu_2$ | Natural death rate of the adult sub-population |
| $\alpha_0$ | Maximum death rate due to AIDS. $(\alpha_i \leq \alpha_0), i = 1, 2$ |
| $\alpha_1$ | Death rate of infected juvenile sub-population due to AIDS |
| $\alpha_2$ | Death rate of infected adult sub-population due to AIDS |
| $\varphi_1$ | Disease induced death rate of infected juveniles not receiving HAART |
| $\varphi_2$ | Disease induced death rate of infected adults not receiving HAART |
| $\varpi_1$ | Disease induced death rate of infected juveniles receiving HAART |
| $\varpi_2$ | Disease induced death rate of infected adults receiving HAART |
| $\tau_1$ | Disease induced death rate of recovered juvenile not receiving HAART |
| $\tau_2$ | Disease induced death rate of removed adults not receiving HAART |
| $T$ | Maximum lifespan after infection ($T = 10$ years) |
| $k$ | Efficacy of HAART ($0 \leq k \leq 1$) |
| $c$ | Average number of sexual partners of adult members of class $I_2$ |
| $c'$ | Average number of sexual partners of adult members of class $R_2$ |
| $\beta$ | Probability of transmission by adult members of class $I_2$ |
| $\beta'$ | Probability of transmission by adult members of class $R_2$ |
| $\rho_1$ | Probability of secondary infection by recovered juveniles population in U=U |
| $\rho_2$ | Probability of secondary infection by recovered adults population in U=U |
| $\sigma_1$ | Proportion of infected juveniles receiving HAART per unit time (Treatment rate) |
| $\sigma_2$ | Proportion of infected adults receiving HAART per unit time (Treatment rate) |
| $\pi_1$ | Proportion of juvenile population from susceptible to exposed/latent class |
| $\pi_2$ | Proportion of adult population from susceptible to exposed/latent class |
| $\varepsilon_1$ | Proportion of removed juveniles still receiving treatment and being moved to susceptible class |
| $\varepsilon_2$ | Proportion of removed adults still receiving treatment and being moved to susceptible class |
| $1 - \xi$ | Proportion of healthy newborns from infected mothers |
| $\xi$ | Proportion of infected newborns from infected mothers |
| $B_1(t)$ | Incidence rate in the juvenile sub-population. $B_1(t) = 0$ (no sexual contact) |
| $B_2(t)$ | Incidence rate or force of infection in the adult sub-population |
| $\eta_j$ | Maturation rate from Juveniles to adults (where $j = s$ denotes the susceptible population, $j = e$ denotes the exposed/latent population, $j = i$ denotes the infected population, $j = r$ denotes the removed population and $j = u$ denotes the undetectable=untransmittable population) |
| $m$ | Fixed ratio of adults to juveniles, $m = \frac{N_2}{N_1}$ |

### 2.2 MODEL ASSUMPTIONS

The following assumptions would help in the derivation of the model:

1. There is no emigration from the total population and there is no immigration into the population.
2. Maturation (or maturity) is interpreted as growth from Juvenile stage into adult.
3. The susceptible population are first exposed to a latent class where they can infected or not.
4. Some infected individuals move to the removed class when counseled and are placed under highly active antiretroviral therapy (HAART).
5. Newborns are not of the same class as their progenitor. A fraction (1 − *ξ*) of newborns from infected mothers are healthy, while the remaining fractions *ξ* are born with the virus.
6. The AIDS cases have full-blown symptoms and are therefore not sexually active.
7. The recruitment into the *S*-class is only through birth for the juvenile sub-population and through maturation for the adult sub-population.
8. The recruitment from the *S*-class into the *E*-class is through birth for newborns and through heterosexual activities for adults. This is done at a rate *π*_1_ and *π*_2_ for the juvenile and adult sub-population respectively.
9. The recruitment into the *R*-class from the *I*-class depends on the effectiveness of public campaign and counselling. This is done at a rate *σ*_1_ and *σ*_2_ for the juvenile and adult sub-populations, respectively.
10. The recruitment into the *U*-class from the *R*-class depends on the effectiveness of the HAART and the change in social behavior of the recovered population. This is done at a rate *ρ*_1_ and *ρ*_2_ for the recovered juvenile and adult sub-population respectively.
11. The recruitment into the *S*-class over again from the *U*-class depends on how long the population in the *U*-class remain in the class while actively receiving treatment. This stage it is assumed that the compartment is filled with fully removed population whose viral load is less than 1% and have 0% chance of secondary infection. This is done at a rate *ε*_1_ and *ε*_2_ for juvenile and adult sub-population respectively.
12. There is a chance of infection by the juvenile and adult population in the U=U class at *ρ*_1_ and *ρ*_2_ probability if the administration of HAART is discontinued at any given time.
13. There is a fixed ratio of adults to juveniles given by 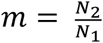. This assumption allow for a suitable control of the population at a given time.

### 2.3 MODEL DESCRIPTION

The susceptible-exposed-infected-removed-undetectable=untransmissible-susceptible (SEIRUS) model that considers an open age-structured population of juvenile and heterosexual individuals is formulated based on the McKendrick-von-Foerster type two-age-structured SIR model as formulated by [1] in studying the effect of antiretroviral therapy. The population is sub-divided based on demographic structure and epidemiological structure. Under a demographic structure, the population is divided into classes, the juvenile class (0 – 14 years) and the adult class (15 years and above), while under the epidemiological structure of this study is divided into six classes, namely; susceptible (*S*), exposed (E), infected (*I*), removed (*R*), Undetectable=Untransmitable (U=U) and those infected progressing to AIDS (*A*). A susceptible is an individual that is yet to be infected, but is open to infection as he or she interacts with members of the *I*-class. An infected individual is one who has contracted HIV and is at some stage of infection. A removed individual is one that is confirmed to be HIV positive, counseled, and is receiving treatment via highly active antiretroviral therapy (HAART). A member of the Undetectable=Untransmitable class is one that has been removed and has been actively receiving treatment through HAART and has been satisfied by the UN-MDG 6’s standard to be in the U=U class. A member of the *A*-class is an individual who is HIV positive, and has progressed to full blown AIDS [3].

The following diagram describes the dynamic of SEIRUS framework, and will be useful in the formulation of model equations.

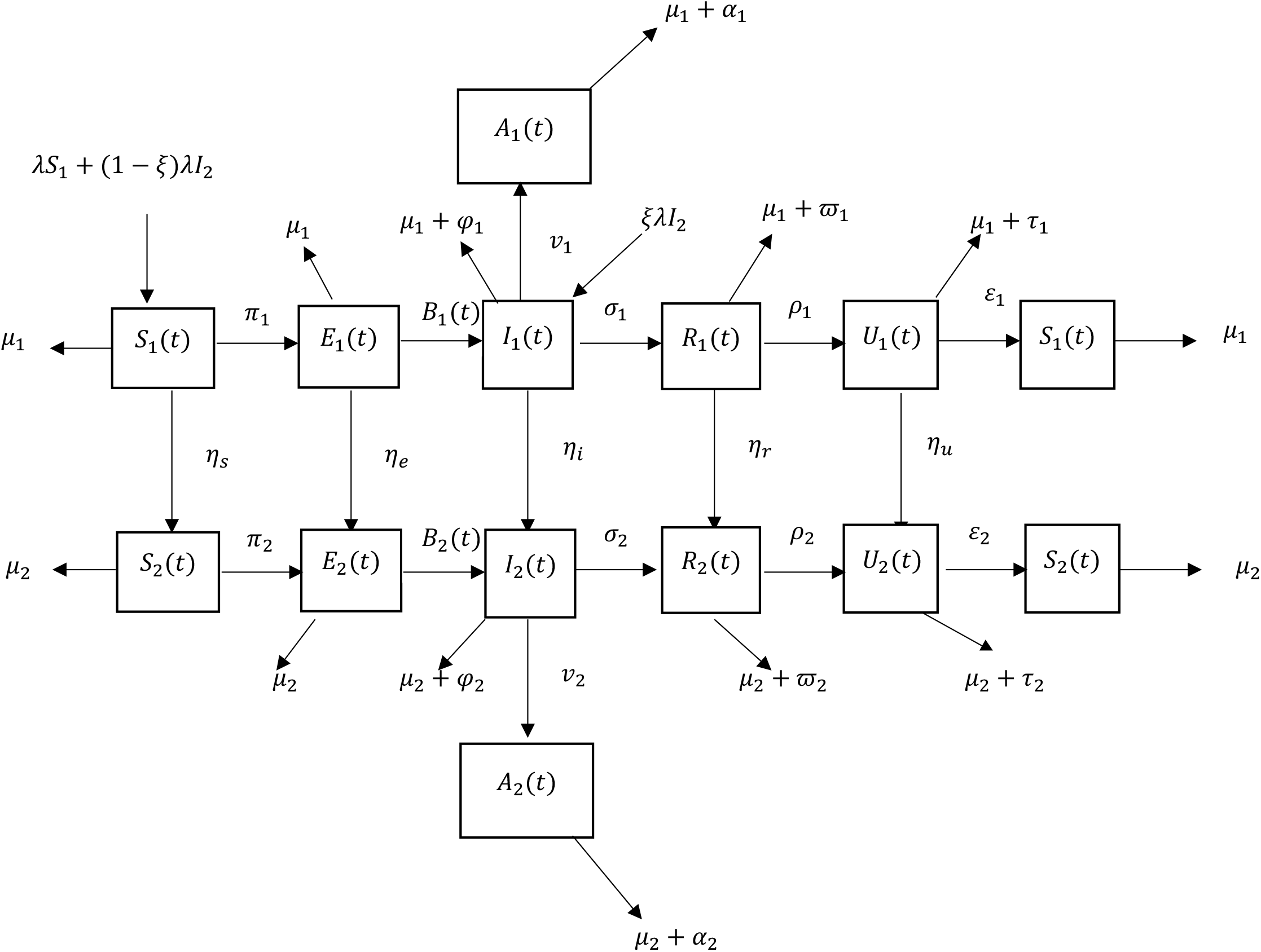

## 3. THE MODEL EQUATIONS

From the assumptions and the flow diagram above, the following model equations are derived.

**For the Juvenile sub-populations:**

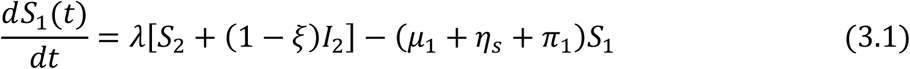

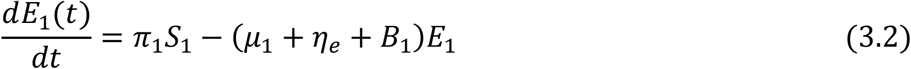

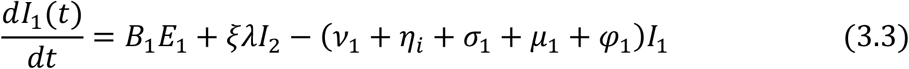

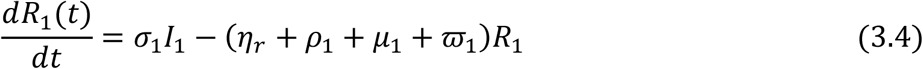

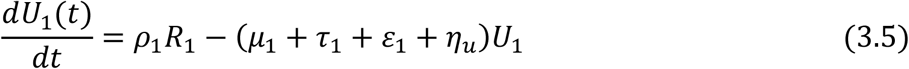

**For the Adult sub-populations:**

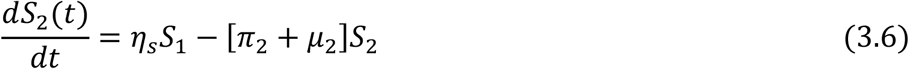

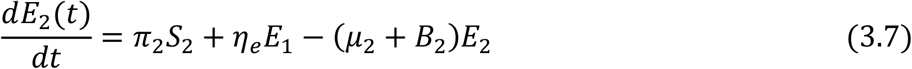

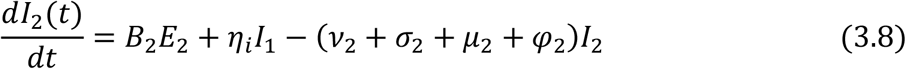

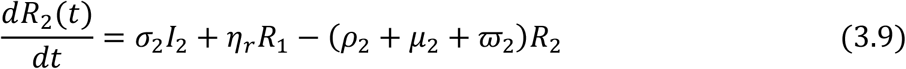

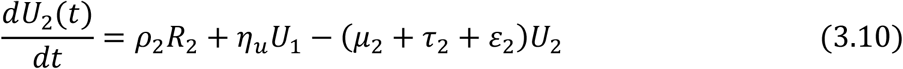

**For those progressing to AIDS**

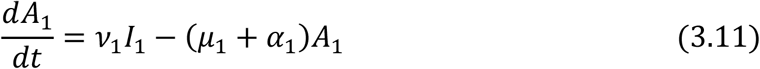

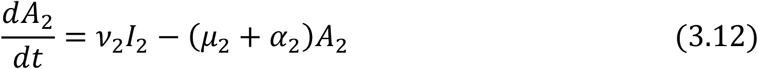

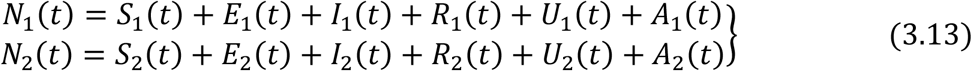

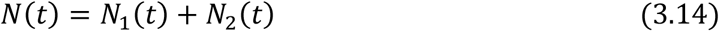

The incidence rate or force of infection at time *t* denoted by *B*_2_(*t*) in the adult population is given as

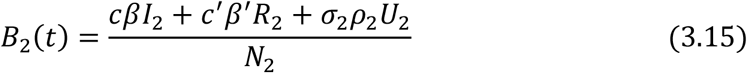

### 3.1 MODEL EQUATIONS IN PROPORTIONS

To simplify the model, it is reasonable to assume that infected juvenile and adult who progress to full blown AIDS are isolated and sexually inactive; hence they are not capable of producing children (vertical transmission) and they do not contribute to viral transmission horizontally (from adult to adult) [4].

To achieve this, we normalize the model by transforming the model equations into proportions and eliminate the AIDS class *A*(*t*), which invariably reduces the number of model equations from twelve to ten. The derive model equations in proportion of infected juveniles and adults define prevalence of infection, which has biological meaning.

The model equations are transformed into proportions as follows;

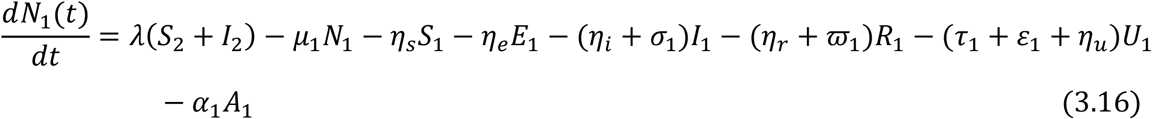

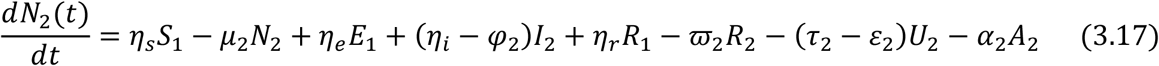

Let

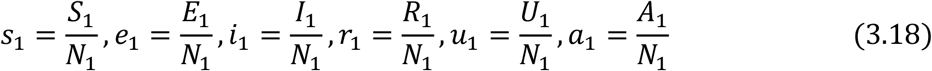

similarly,

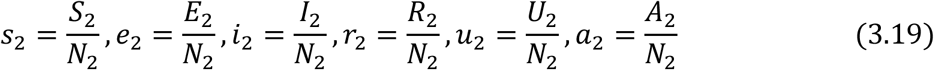

and

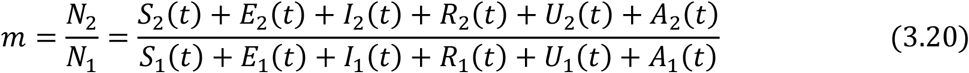

Then the normalized system is follows,

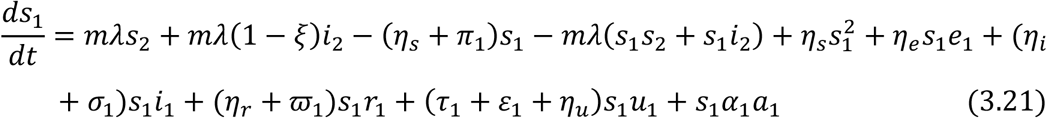

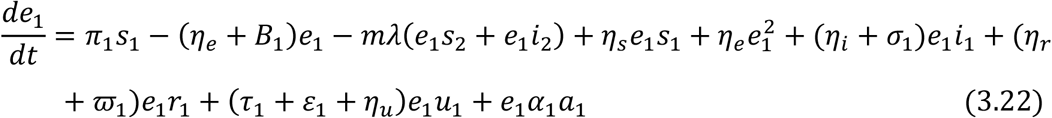

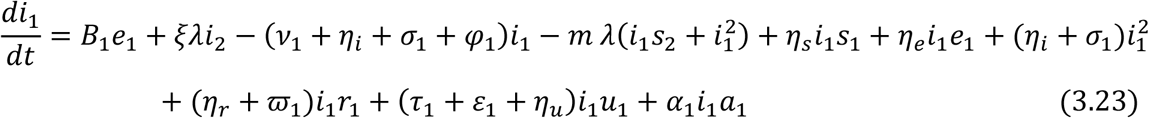

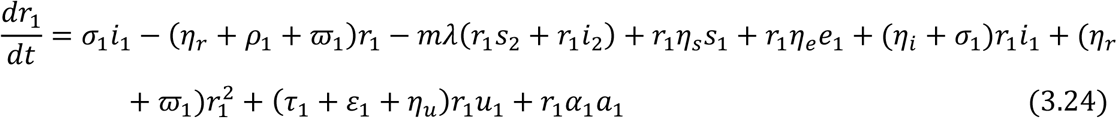

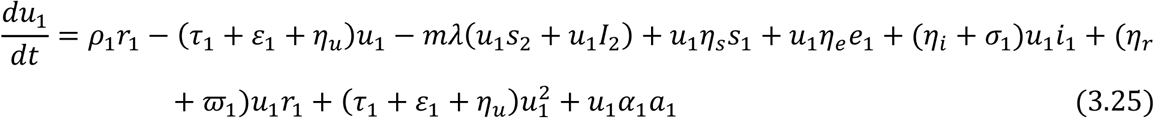

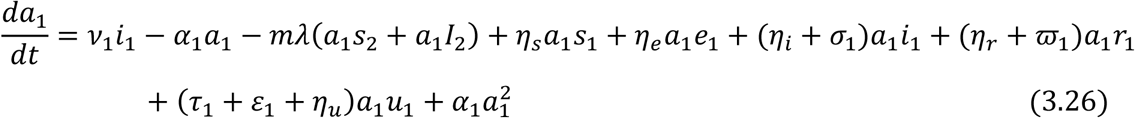

Similarly for the adult sub-population, the normalized system is follows:

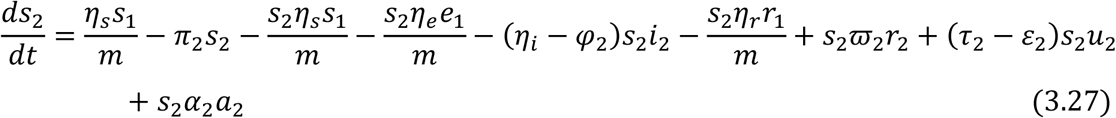

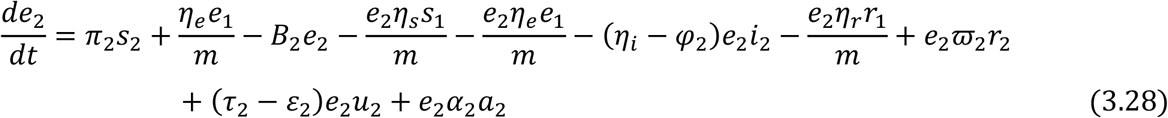

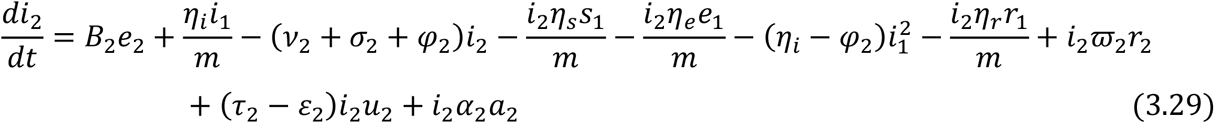

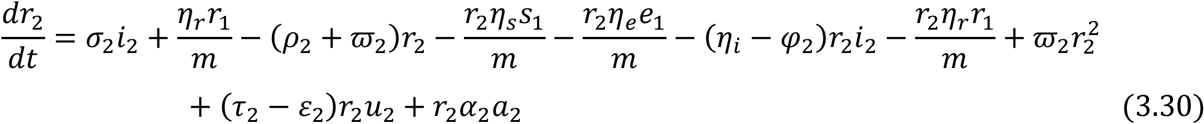

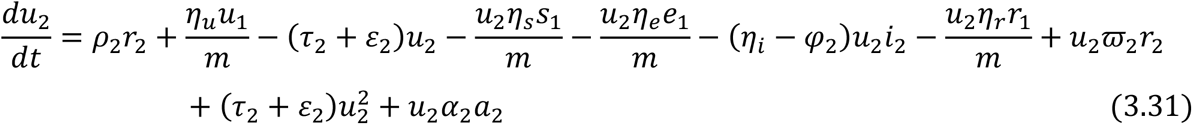

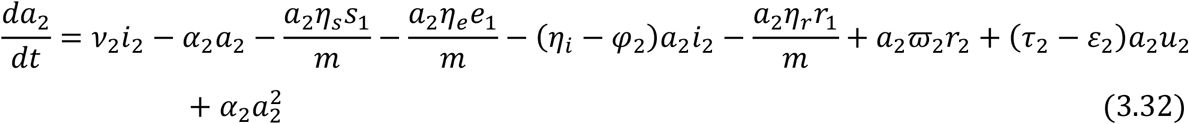

However,

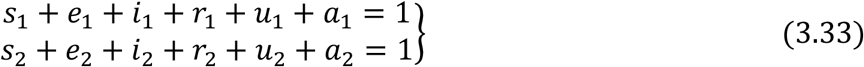

Gives the following governing equations of the model below:

For the Juvenile sub-population:

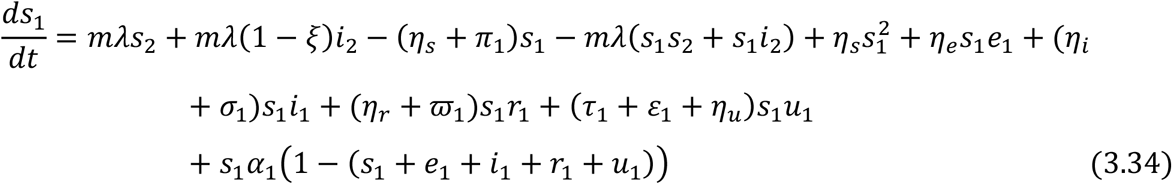

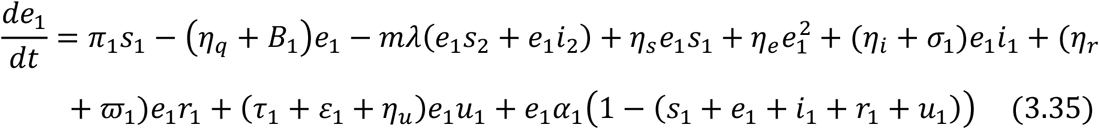

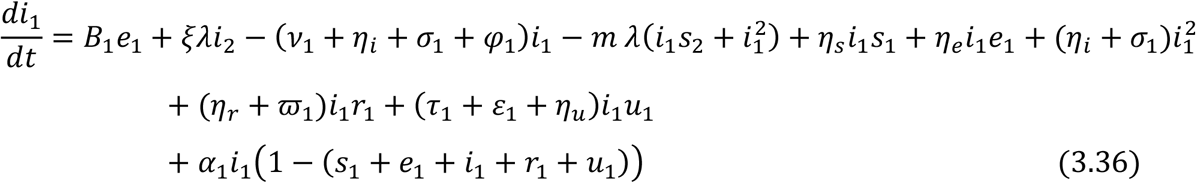

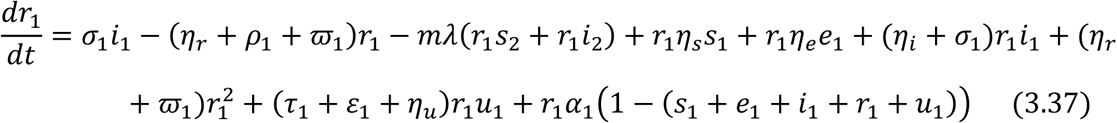

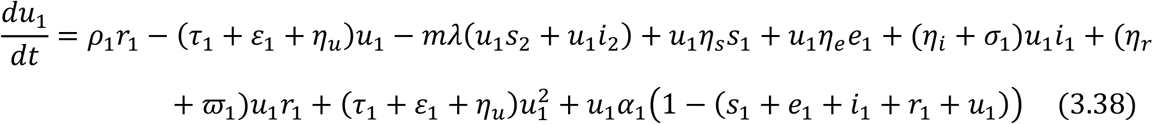

For the Adult sub-population

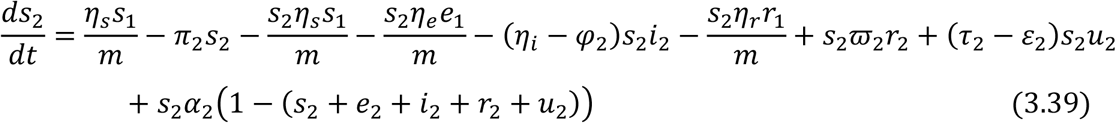

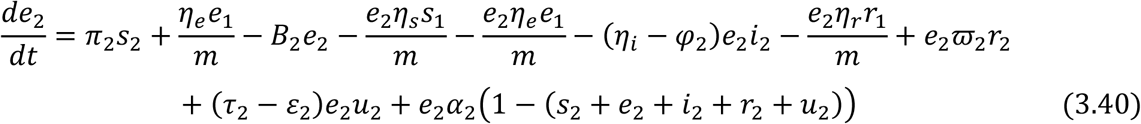

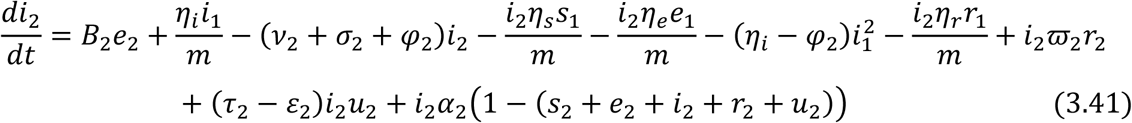

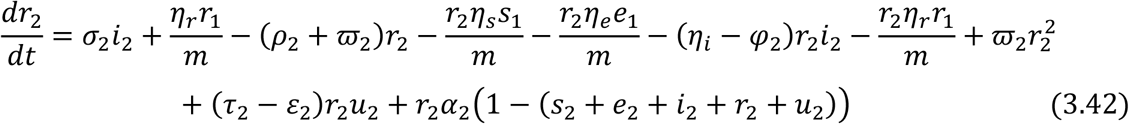

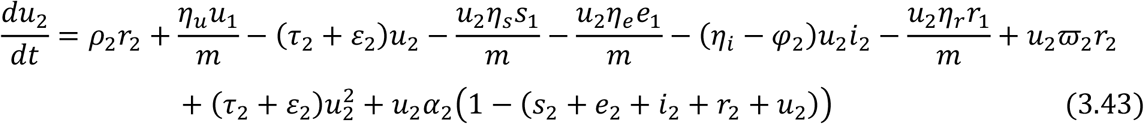

Equations (3.34) to (3.43) are the model equations in proportions, which define prevalence of infection.

### 3.2 EXISTENCE AND UNIQUENESS OF DISEASE FREE EQUILIBRIUM STATE (*E*_0_) OF THE SEIRUS MODEL

The disease-free equilibrium (DFE) state of the endemic SEIRUS model is obtained by setting the left hand sides of equations (3.34) – (3.43) to zero while setting the disease components *e*_1_ = 0, *e*_2_ = 0, *i*_1_ = 0, *i*_2_ = 0, *r*_1_ = 0, *r*_2_ = 0 and *u*_1_ = 0, *u*_2_ = 0 leading to equations (44) – (45) below

For the Juvenile sub-population:

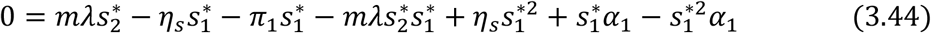

For the Adult sub-population:

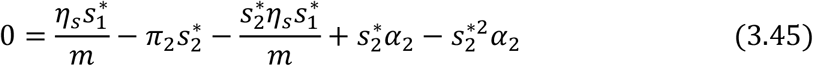

Factorizing 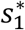 From Equation (3.45) and substituting into (3.44) gives;

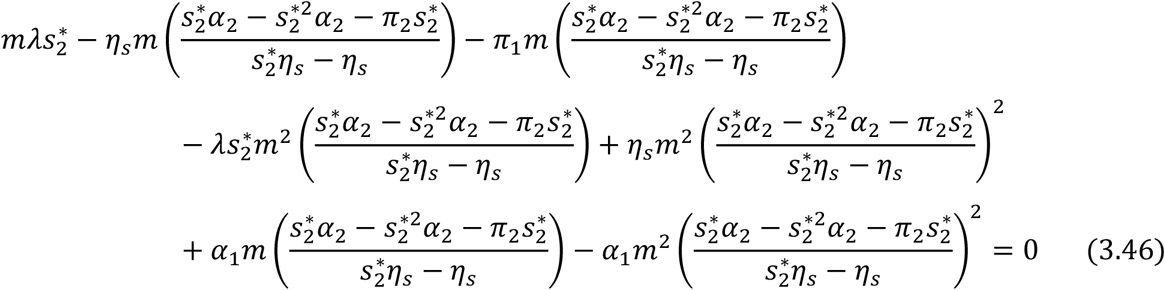

Multiplying through by 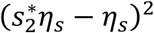, gives

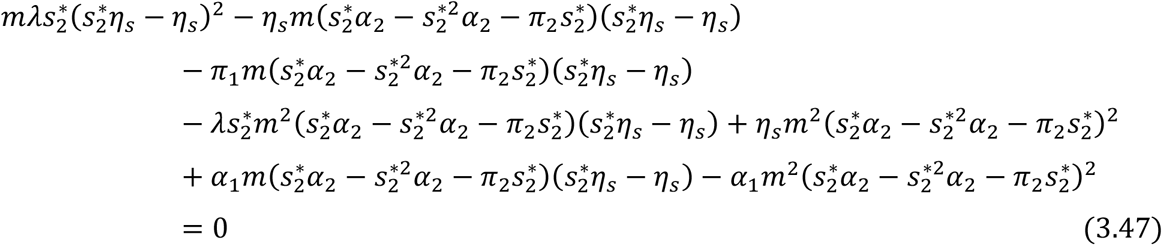

Simplifying further and collecting like terms in 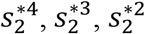 and 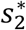 gives,

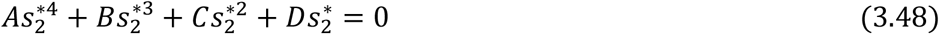

where

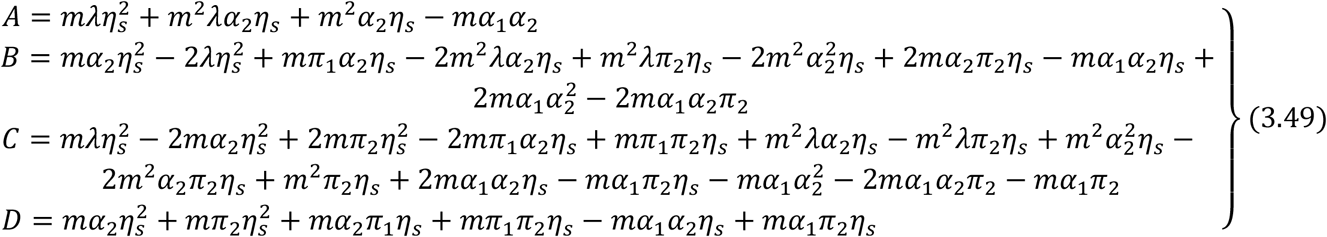

Therefore, the solution for the simultaneous equations (3.48) is given by

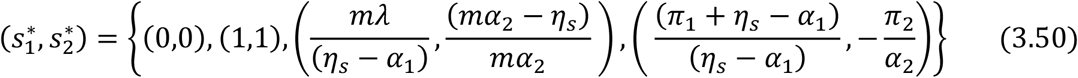

Ignoring the native values of 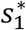 and 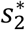 and other stringent conditions, there exist a unique trivial and disease-free equilibrium states at 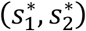 given by (0,0) and (1,1) respectively.

The solution (3.50) satisfies equations (3.47) identically.

### 3.3 COMPUTATION OF THE BASIC REPRODUCTIVE NUMBER (*R*_0_) OF THE MODEL

The Basic Reproductive number (*R*_0_) is define as the number of secondary infections that one infectious individual would create over the duration of the infectious period, provided that everyone else is susceptible. *R*_0_ = 1 is a threshold below which the generation of secondary cases is insufficient to maintain the infection in human community. If *R*_0_ < 1, the number of infected individuals will decrease from generation to next and the disease dies out and if *R*_0_ > 1 the number of infected individuals will increase from generation to the next and the disease will persist. To compute the basic reproductive number (*R*_0_) of the model (3.33) – (3.43), we employ the next generation method as applied by [1], [5] and [6].

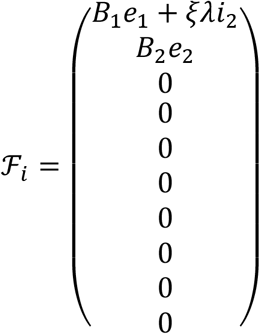

and

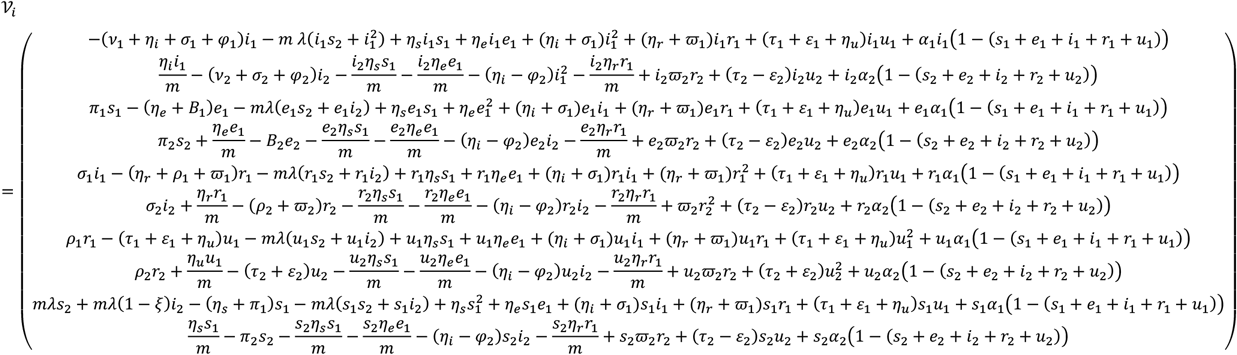

where *ℱ*_*i*_ and *𝒱*_*i*_ are the rate of appearances of new infections in compartment *i* and the transfer of individuals into and out of compartment *i* by all means respectively. Using the linearization method, the associated matrices at disease-fee equilibrium (*E*_0_) and after taking partial derivatives as defined by

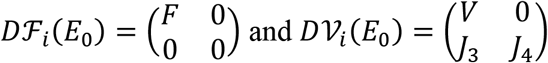

where *F* is nonnegative and *V* is a non-singular matrix, in which both are the *m* × *m* matrices defined by

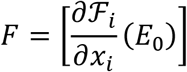

and

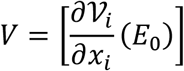

with 1 ≤ *i, j* ≤ *m* and *m* is the number of infected classes.

In particular *m* = 2, we have

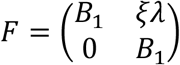

and

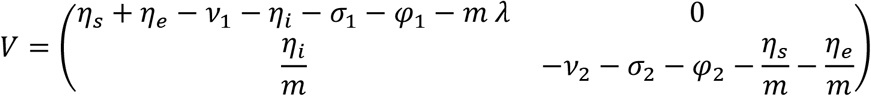

If the inverse of *V* is given as

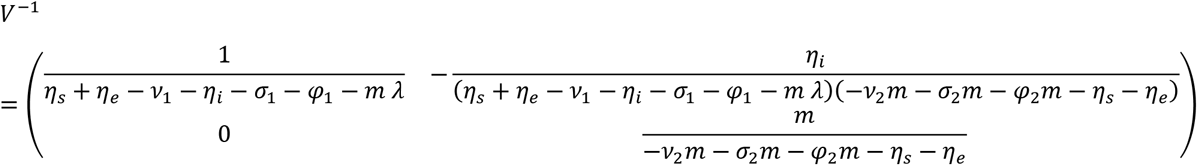

Then the next matrix denoted by *FV*^−1^ is given as

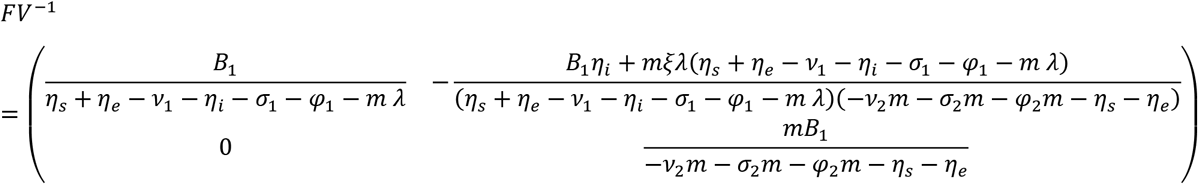

We find the eigenvalues of *FV*^−1^ by setting the determinant |*FV*^−1^ − *γI*| = 0

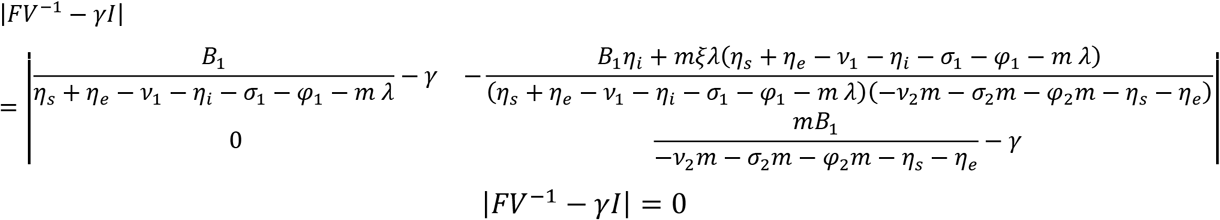

with characteristics polynomial

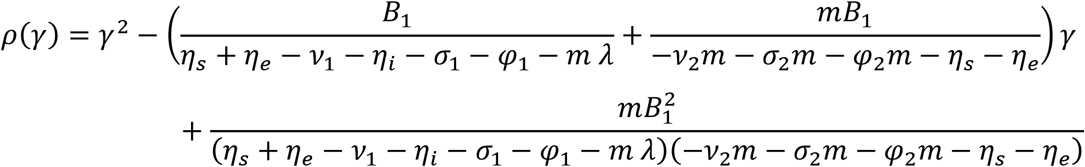

and characteristics equation given as

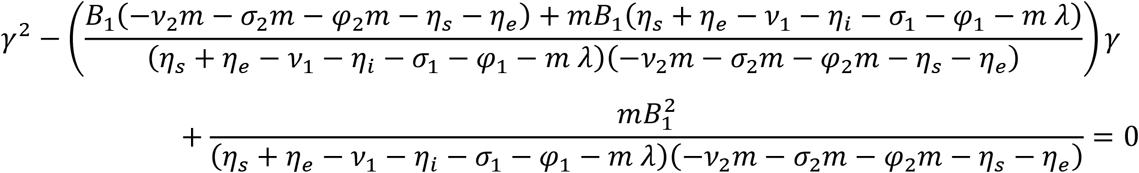

Solving the characteristics equation for the eigenvalues *γ*_1,2_, where *R*_0_ is the maximum of the two eigenvalues *γ*_1,2_. Hence the Basic Reproductive number is the dominant eigenvalues of *FV*^−1^. Thus we have that

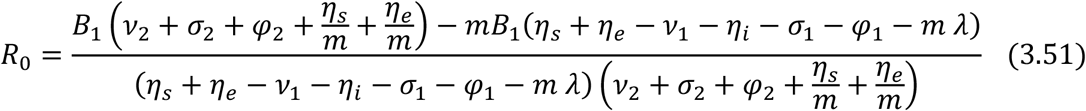

Because the incidence rate in the juvenile population is zero, that is, there is no transmission of the disease between children to children, and *B*_1_(*t*) = 0 from table 1, then equation (3.51) hence

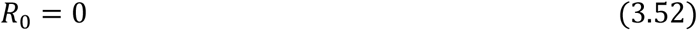

**Table 1.** Estimated values of the parameters used in the Numerical experiments.

| Parameters | Values | Source | Parameters | Values | Source |
| --- | --- | --- | --- | --- | --- |
| $N(0)$ | 203,248,775 | [9] | $c'$ | 1.00** | Assumed |
| $N_1(0)$ | 85,364,485 | [9] | $B_2(t)$ | 0.01037146* | Author's computation |
| $N_2(0)$ | 117,884,290 | [9] | $T$ | 10 | [8] |
| $s_1(0)$ | 0.41999999 | [9] | $\alpha_0$ | 0.0408 | [8] |
| $e_1(0)$ | 0.42 | [9] | $\alpha_1$ | 0.0144 | [9] |
| $i_1(0)$ | 0.0164 | [9] | $\rho_1$ | 0.012 | [9] |
| $r_1(0)$ | 0.007799 | [9] | $\alpha_2$ | 0.0122 | [9] |
| $u_1(0)$ | 0.00073 | [9] | $\varphi_1$ | 0.0760 | [9] |
| $v_1$ | 0.125 | [11] | $\rho_2$ | 0.35 | [9] |
| $s_2(0)$ | 0.579 | [9] | $\varphi_2$ | 0.0520 | [9] |
| $e_2(0)$ | 0.58 | [9] | $\mu_1$ | 0.0890 | [9] |
| $i_2(0)$ | 0.0149 | [9] | $\pi_1$ | 0.025** | Assumed |
| $r_2(0)$ | 0.0088646 | [9] | $\mu_2$ | 0.01512 | [9] |
| $u_2(0)$ | 0.00083 | [9] | $\lambda$ | 0.038098 | [8] |
| $v_2$ | 0.070 | [11] | $\pi_2$ | 0.05** | Assumed |
| $\eta_s$ | 0.05*** | Assumed | $m$ | 1.3809524 | [9] |
| $\eta_e$ | 0.04*** | Assumed | $K$ | 0.5 | [9] |
| $\eta_i$ | 0.03*** | Assumed | $\beta$ | 0.150 | Assumed |
| $\eta_r$ | 0.02*** | Assumed | $\beta'$ | 0.010 | Assumed |
| $\eta_u$ | 0.01*** | Assumed | $\sigma_1$ | 0.25** | Assumed |
| $\varpi_1$ | 0.0005121* | Computed | $\sigma_2$ | 0.25** | Assumed |
| $\varpi_2$ | 0.0003503* | Computed | $1 - \xi$ | 0.85** | Assumed |
| $c$ | 1.00** | Assumed | $\xi$ | 0.15** | Assumed |
| $\varepsilon_1$ | 0.0234504 | Assumed | $\tau_1$ | 0.05432 | Assumed |
| $\varepsilon_2$ | 0.15120 | Assumed | $\tau_2$ | 0.19854 | Assumed |
| $a_1$ | 0.0048 | Computed | $a_2$ | 0.005037 | Computed |

**Computed based on parameter values\***: $\varpi_i = \varphi_i e^{-kT} B_2(t) = (c\beta I_2 + c'\beta' R_2 + \sigma_2 \pi_2 E_2)/N_2$
**Assumed\*\***: Hypothetical data use for research purpose (See section 4.4.1) **Assumed\*\*\***: Based on [1].

The Basic Reproductive number (*R*_0_) by Equation (3.52) shows that the number of secondary infections that one infectious individual would create over the duration of the infectious period, provided that everyone else is susceptible is zero and this implies there is no secondary infection in an endemic situation.

## 4. DESCRIPTION AND VALIDATION OF BASELINE PARAMETERS

The population of Nigeria as estimated by the [7] was 203,248,775 with males scooping more than half the entire population having a total number of 108,345,123 and the females holding at 94,903,652. It should be noted, though, that while women are slightly outnumbered by men, after the age of 65, women outnumber the number of men. Nigeria’s population is predicted to hit 206 million by 2020 and 264 million by 2030 – crossing 300 million threshold around 2036. Furthermore, with a relatively young country, Nigeria has a young population structure, with almost half of the population being under the age of 15 years. Young people, 15-24 years of age constitute more than a quarter of the population and the median age according to [7], for both males and females of the country actually 18.4 years in 2019.

The life expectancy in Nigeria is, unfortunately, the lowest in all of West Africa. The average life expectancy is around 54.5 years of age according to WHO data [8], with men living an average of 53.7 years and women living an average of 55.4 years. This very low number can be attributed to the fact that the country has a lot of health issues. As previously mentioned, the AIDS epidemic is a major player in the low life expectancy. But on top of that, Nigeria has fallen victim to a high child and maternal mortality rate and the wide spread growth of mother-to-child transmission of HIV as well as polio virus. According to [7], one out of every five children that are born in Nigerian will die before they reach the age of five due to the many health risks in Nigeria.

While pregnancy is obviously not a disease by any means, a lot of expectant mothers in Nigeria die from pregnancy complications every year. A Nigerian woman’s chances of death during pregnancy or childbirth is 1 in 13. However, the components of population change in Nigeria shows that one birth occurs every 4 seconds and one death occurs every 14 seconds. Meanwhile, the population is also affected by the rate of migration into the country as there is one net migrant every 9 minutes into Nigeria which means a net gain of one person every 6 seconds either from immigration or birth and this speaks more about the rapid population growth in the country.

With an estimated population of 203 million people, Nigeria has a total of 1,900,000 people living with HIV as at 2018 and there are 0.65 infections among all people of all ages which are the number of new HIV infections among the uninfected population over a year. The percentage of people living with HIV – among adults (15 – 49 years) was recorded to be 1.5% with a total of 130,000 newly infected adult population and 140 newly juvenile population in 2018 and 53,000 cases of death due to AIDS-related illness.

Although there has been progress in the number of AIDS-related deaths in Nigeria since 2010, with a 26% decrease, from 72,000 deaths to 53,000 deaths. However, the number of new HIV infections has risen, from 120,000 to 130,000 in the same period [9]. With the 90-90-90 targets vision for 2020, which implies a 90% of people living with HIV knowing their HIV status, 90% of people who know their HIV-positive status being able to access treatment and 90% of people being placed on treatment to have suppressed viral loads, it was observed based on the 90-90-90 target that in 2018 only 67% of people living with HIV knew their status and of the 81% of people living with HIV who are supposed to be on treatment, only 53% of them were on treatment and of the 73% of people who were expected to have virally suppressed the disease, only 42% of them have virally suppressed it which shows a sharp contrast from the envisioned target of 2020.

According to UNAID factsheet [9], of all adults aged 15 years and over living with HIV, 55% were on treatment as at 2018 while only 35% of children/juvenile aged 0 – 14 years living with HIV were on treatment. Also, 4% of pregnant women living with HIV accessed antiretroviral medicine to prevent transmission of the virus to their baby (MTC), which has helped to prevent about 7,200 new HIV infections among new-borns. The report further shows that women are disproportionally affected by HIV in Nigeria as of the 1,800,000 adults living with HIV, 1,000,000 (55.6%) were women. Whereby new HIV infections among young women aged 15 – 24 years were less than double those among young men with 26,000 new infections among young women, compared to 15,000 among young men. More so, HIV treatment was found to be higher among women than men, however, with 68% of adult women living with HIV on treatment, compared to 37% of adult men.

The baseline parameters for Nigeria include total population (both juvenile and adult), birth rate **(*λ*)**, natural death rate for juvenile (*μ*_1_) and natural death rate for adult (*μ*_2_) as estimate gotten from [9]. See Table 1 for details.

### 4.1 DESCRIPTION AND VALIDATION OF OTHER ESTIMATED PARAMETERS

#### (a) Proportion of susceptible, exposed, infected, removed and undetectable=untrasmittable individuals in both the Juvenile and Adult sub-population

(*s*_1_, *s*_2_, *e*_1_, *e*_2_, *i*_1_, *i*_2_, *r*_1_, *r*_2_, *u*_1_, *and u*_2_)

The United Nations AIDS [9] and the World Population Review [7] estimates there are about was 1,900,000 people living with HIV in Nigeria, ranking the country with the second highest burden of HIV infection in Africa after South Africa and the World at large followed by Mozambique and India respectively. Females constitute over half (55.56%) of people living with HIV in Nigeria, making a total of about 1,000,000 women and girls are infected with HIV and a total of 900,000 males. The estimated overall HIV prevalence rate is approximately 1.4% among the Nigerian Population. For adults aged between 15 – 49 years, an estimated 1.5% of the population is HIV positive.

For the purpose of this study we use 203,248,775 as the estimated population for Nigeria, with a total of 85,364,485 juveniles (0 – 14 years) and 117,884,290 adults (15 years and above) according World Population Review (2019) updates. This put the proportion of susceptible juveniles and adult to be *s*_1_(0) = 0.41999999 and *s*_2_(0) = 0.58 respectively.

However, in this study we take the proportion of susceptible population to also start for the proportion of exposed with the assumption that all the susceptible class are equally vulnerable to be exposed to being infected in an ideal situation where MTC and adult behaviours are left unsupervised. Therefore, this put the proportion of exposed juvenile and adult to be *e*_1_(0) = 0.42 and *e*_2_(0) = 0.58 respectively.

Current estimate according to [9] put the number of infected juveniles and adult in South Africa to be about 1,900,000, with a total of 140,000 juveniles and 1,760,000 adults. Using similar approach as in the susceptible compartment where *N*_1_ = 85,364,485 juveniles (0 – 14 years) and *N*_2_ = 117,884,290 adults (15 years and above), we therefore have the proportion of infective to be *i*_1_(0) = 0.0164 for the juveniles and *i*_2_(0) = 0.0149 for adult populations.

According to UNAID factsheet [9], about 53% of people living with HIV in Nigeria who need Highly Active Antiretroviral Treatment (HAART) have access to it making a total of about 1,007,000 people having assess to HAART in 2018 with about 665,800 juveniles and 1,045,000 adults where *N*_1_ = 85,364,485 juveniles (0 – 14 years) and *N*_2_ = 117,884,290 adults (15 years and above). We therefore have the proportion of infective receiving treatment in the juvenile and adult sub-population as *r*_1_(0) = 0.0077995 and *r*_2_(0) = 0.0088646 respectively.

Also, with a success rating in achieving the 90-90-90 targets envision for 2020, 67% of people living with HIV in Nigeria knew their status and need HAART and 53% of them being actively under treatment of HAART. As a result of that about 42% of the 1,007,000 having assess to HAART in 2018 were virally suppressed, that is, the HIV in the 422,940 people is undetectable and the HIV treatment brings the level of the HIV in the body to such a low level that tests cannot detect it. As long as the HAART is adhered to and viral load remains undetectable (and monitored) they remain untrasmitable and hence cannot transmit to others and their health is not affected by HIV. Therefore, according to [9] 53% of the Nigerian infective population (55% and 55% adults and juvenile respectively) were on HAART, hence, 42% of that 55% adults and 35% juvenile are undetectable and untransmitable which makes up a total of 97,699 and 62,172 undetectable=untransmitable adult and juvenile sub-populations respectively with *N*_1_ = 85,364,485 juveniles (0 – 14 years) and *N*_2_ = 117,884,290 adults (15 years and above). We therefore have the proportion of virally suppressed population in the juvenile and adult sub-population as *u*_1_(0) = 0.00073 and *u*_2_(0) = 0.00083 respectively.

#### (b) Death rate due to AIDS (*α*_1_, *α*_2_) and maximum death rate due to AID (*α*_0_)

These are all gotten from [9]. See Table 1.

#### (c) Disease induced death rate of the infected juveniles and infected adults not receiving HAART (*φ*_1_, *φ*_2_)

These are all gotten from [9]. See Table 1.

#### (d) Disease induced death rate of the infected juveniles and infected adults (*ϖ*_1_, *ϖ*_2_) receiving HAART

To get the value for *ϖ*_1_ and *ϖ*_2_, we use the formula *ϖ*_*i*_ = *φ*_*i*_*e*^−*kT*^ where *i* = 1, 2 represent the juvenile and adult sub-population respectively. *ϖ*_1_ and *ϖ*_2_ represents the death rate of the juvenile and adult sub-population who are not receiving HAART. *k* is the efficacy of the drug and *T* is the maximum lifespan after infection, as provided by [10]. See Table 1.

#### (e) The rate of progression from HIV to AIDS in the juvenile and adult sub-population (*v*_1_, *v*_2_)

Without loss of generality, the rate of progression from HIV to AIDS in the juvenile and adult sub-population is taken to be *v*_1_ = 0.125 and *v*_2_ = 0.070 as early diagnosis of HIV infection in children is essential because due to weaker immune systems, the infection in infants and children tends to progress faster than in adults.

#### (f) Maturation rate of susceptible, exposed, infected, removed and undetectable juvenile to adults (*η*_*s*_, *η*_*e*_, *η*_*i*_, *η*_*r*_ and *η*_*u*_)

Maturation is achieved by transferring a portion of the susceptible juvenile to its corresponding susceptible adult sub-population. In the susceptible juvenile compartment, we estimate the number of children alive, for each distinct age between 0 and 14, based on the annual mortality and population growth rate of Nigeria. We then divide the number of 14 years old by the total size of the juvenile sub-population [1]. This will result in the rate of children who will turn 13 and will thus enter the sexually active adult class. The maturation rate for susceptible is thought to be higher than that for the infected population, which in turn is higher for the removed class receiving treatment. In the current research work, the estimated value for the maturation rate for each compartment is given as *η*_*s*_ = 0.05, *η*_*e*_ = 0.04, *η*_*i*_ = 0.03, *η*_*r*_ = 0.02 and *η*_*u*_ = 0.01.

#### (g) Probability of transmission by adult members of class *I*_2_ and class *R*_2_ (*β, β*′)

The term *β* and *β*′ are referred to as probabilistic terms that lies between 0 and 1 and it is expected that *β*′ < *β*. In this research work, we choose to adopt probability of transmission values from [1] which states that probability of transmission is low if it falls within the range (*β* ≤ 0.015, *β*^′^ ≤ 0.00136) and it is high when it falls within the range (*β* ≥ 0.150, *β*^′^ ≥ 0.010). For example in every 1000 adults, 15 transmit the disease in the infected compartment and 1 transmit the disease in the removed compartment is regarded as low transmission rate. Similarly, in every 1000 adults, 150 transmit the disease in the infected compartment and about 10 transmit the disease in the removed compartment is regarded as high transmission rate.

#### (h) Probability of secondary infection by recovered juveniles and adult populations in *U* = *U* compartment (*ρ*_1_, *ρ*_2_)

The recruitment into the *U*-class from the *R*-class depends on the effectiveness of the HAART and the change in social behavior of the recovered population. This is done at a rate *ρ*_1_ and *ρ*_2_ for the recovered juvenile and adult sub-population respectively.

The recruitment into the *S*-class over again from the *U*-class depends on how long the population in the *U*-class remain in the class while actively receiving treatment. This stage it is assumed that the compartment is filled with fully removed population whose viral load is less than 1% and have 0% chance of secondary infection. This is done at a rate *ε*_1_ and *ε*_2_ for juvenile and adult sub-population respectively.

There is a chance of infection by the juvenile and adult population in the U=U class at *ρ*_1_ and *ρ*_2_ probability if the administration of HAART is discontinued at any given time.

The term *ρ*_1_ and *ρ*_2_ are referred to as probabilistic terms that lies between 0 and 1 and it is expected that *ρ*_1_ < *ρ*_2_ the probability of re-infection by juvenile is almost negligible but for the purpose of accuracy we take *ρ*_1_into consideration no matter how small. In this research work, probability of transmission is low if it falls within the range (*ρ*_2_ ≤ 0.25, *ρ*_1_ ≤ 0.0016) and it is high when it falls within the range (*ρ*_2_ ≥ 0.35, *ρ*_1_ ≥ 0.012).

#### (i) Treatment rate of the juvenile and adult sub-population (*σ*_1_, *σ*_2_)

The term *σ*_1_ and *σ*_2_ are referred to as the proportion of those receiving treatment in comparison with the juvenile and adult sub-population respectively. It expressed as 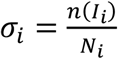. The treatment rate is low when it falls within the range (*σ*_1_ ≤ 0.25, *σ*_2_ ≤ 0.25) for the juvenile and adult sub-population. Similarly treatment rate is high when it falls within the range (*σ*_1_ ≥ 0.85, *σ*_2_ ≥ 0.85). For example in every 100 juveniles or adults that are infected, when 25 or less receive treatment, then it is regarded as low treatment rate, while in every 100 juveniles or adults, when 75 and above receive treatment, it is regarded as high treatment rate.

#### (j) Rate of exposure or latency rate of juvenile and adult sub-population (*π*_1_, *π*_2_)

The recruitment from the *S*-class into the *E*-class is through birth for newborns and through heterosexual activities for adults. This is done at a rate *π*_1_ and *π*_2_ for the juvenile and adult sub-population respectively. Due to the care given to pregnant mothers and proper vaccination during pregnancy the rate of latency for juvenile will be low as compared to that of adults. For adults, a lot of factors exposes them or makes them more latent than the juvenile and some of this factors could include heterosexual relationships, use of unsterilized syrings for drugs and medication, and other uncultured behaviours hence in this research, *π*_1_ < *π*_2_.

#### (k) Average number of sexual partners in the *I*_2_ and *R*_2_ class (*c, c*^′^)

The average number of sexual partners in the infected class and removed class is 1 respectively. Although it is expected that *c*^′^ < *c* since *β*^′^ < *β*.

#### (l) Proportion of infected newborn (*ξ*) and healthy (1 − *ξ*) newborn

The term *ξ* and (1 − *ξ*) are referred to as the proportion of those children born with the disease and those born healthy. Hence this parameter must lies between 0 and 1 (0 ≤ *ξ* ≤ 1). It expressed as

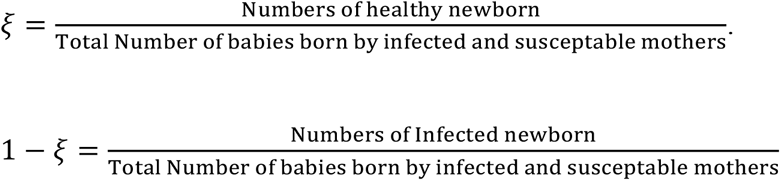

#### (m) Incidence rate in juvenile and adult sub-population (*B*_1_(*t*), *B*_2_(*t*))

The incidence rate in the juvenile sub-population is negligible because there is no sexual contact and hence *B*_1_(*t*) = 0 and that of the adult sub-population, because there is actively a force of infection, *B*_2_(*t*) > 0.

#### (n) Probability of induced death of juvenile and adult sub-population not receiving HAART (*τ*_1_, *τ*_2_)

These are all gotten from [9]. See Table 1.

#### (o) Proportion of removed juveniles and adult still receiving treatment and being moved to susceptible class (*ε*_1_, *ε*_2_)

These are all gotten from [9]. See Table 1.

### 4.2 NUMERICAL EXPERIMENTS OF THE MODEL

The age-structured deterministic model (3.34) – (3.43) was solved numerical using Runge-Kutta-Fehllberg 4-5th order method and implemented using Maple 15 Software (Maplesoft, Waterloo Maple Inc, 2012). The model equations were first transformed into proportions, thus reducing the model equations to ten differential equations. The parameters used in the implementation of the model are shown in Table 1 below. Parameters were chosen in consonance with the threshold values obtained in the stability analysis of the disease free equilibrium state of the model.

### 4.3 GRAPHICAL REPRESENTATION OF RESULTS

Experiment 1**: The effect of treatment on recovery in the juvenile sub-population when the probability of secondary transmission is low (*ρ***_**1**_ = **0. 012, *ρ***_**2**_ = **0. 35) in the adult sub-population**

Experiment 2**: The effect of treatment on recovery in the adult sub-population when the probability of secondary transmission is low (*ρ***_**1**_ = **0. 012, *ρ***_**2**_ = **0. 35) in the adult sub-population**

Experiment 3**: The effect of treatment on recovery in the juvenile sub-population when the probability of secondary transmission is high (*ρ***_**1**_ = **0. 150, *ρ***_**2**_ = **0. 010) in the adult sub-population**

Experiment 4**: The effect of low treatment rate (*σ***_**1**_ = ***σ***_**2**_ ≤ **0. 25) recovery in the juvenile and adult**

**sub-population when the probability of secondary transmission is low (*ρ***_**1**_ = **0. 012, *ρ***_**2**_ = **0. 35)**

Experiment 5**: The effect of high treatment rate (*σ***_**1**_ = ***σ***_**2**_ ≥ **0. 85) on recovery in the juvenile and adult sub-population when the probability of secondary transmission is low (*ρ***_**1**_ = **0. 012, *ρ***_**2**_ = **0. 35)**

Experiment 6**: The effect of low treatment rate (*σ***_**1**_ = ***σ***_**2**_ ≤ **0. 25) on recovery in the juvenile and adult sub-population when the probability of secondary transmission is high (*ρ***_**1**_ = **0. 150, *ρ***_**2**_ = **0. 010)**

Experiment 7**: The effect of high treatment rate (*σ***_**1**_ = ***σ***_**2**_ ≥ **0. 85) on recovery in the juvenile and adult sub-population when the probability of secondary transmission is high (*ρ***_**1**_ = **0. 150, *ρ***_**2**_ = **0. 010)**

Experiment 8**: The effect of high treatment rate (*σ***_**2**_ ≥ **0. 85) on recovery when the probability of secondary transmission is low (*ρ***_**1**_ = **0. 012, *ρ***_**2**_ = **0. 35) and the juvenile sub-population is left untreated**.

Experiment 9**: The effect of high treatment rate (*σ***_**2**_ ≥ **0. 85) on recovery when the probability of secondary transmission is high (*ρ***_**1**_ = **0. 150, *ρ***_**2**_ = **0. 010) and the juvenile sub-population is left untreated**.

Experiment 10**: The effect of low treatment rate (*σ***_**1**_ ≤ **0. 25) on recovery when the probability of secondary transmission is low (*ρ***_**1**_ = **0. 012, *ρ***_**2**_ = **0. 35) and the adult sub-population is left untreated**

Experiment 11**: The effect of high treatment rate (*σ***_**1**_ ≥ **0. 85) on recovery when the probability of secondary transmission is high (*ρ***_**1**_ = **0. 150, *ρ***_**2**_ = **0. 010) and the adult sub-population is left untreated**

Experiment 10**: The effect of low treatment rate (*σ***_**1**_ = ***σ***_**2**_ ≤ **0. 25) on recovery in the juvenile and adult sub-population when all newborns from infected mothers are HIV positive** (***ξ*** = **1. 0**)

Experiment 13**: The effect of high treatment rate (*σ***_**1**_ = ***σ***_**2**_ ≥ **0. 85) on recovery in the juvenile and adult sub-population when all newborns from infected mothers are HIV positive**(***ξ*** = **1. 0**)

Experiment 15**: The effect of vertical transmission on the recovery in the juvenile and adult sub-population when newborns from infected mothers are HIV positive at different proportion ( *ξ***_+_ = **1. 0, *ξ***_−_ = **0. 0)**

### 4.4 DISCUSSION OF RESULTS

In experiment 1 and 3, the effect of HAART treatment on the recovery compartment to the undetectable=untransmitable compartment in the juvenile sub-population was investigated as shown in Figure 4.1 and Figure 4.3 when the probability of secondary transmission is low and high respectively. From Figure 4.1 as the probability of secondary transmission in the adult sub-population seems to decline progressively (*σ*_1_ = 0.25) due to the administration of HAART to the recovered compartment, the recovery rate in the juvenile sub-population is seen to rise progressively in a selected period of time while the rate of recovery for in adult sub-population seems to rise in a short time and eventually drops over the said period of time. It therefore means if the treatment is administered progressively mostly among women and there is less MTC vertical transmission, there will continually be a stiff rise in the recovery rate among juvenile as the secondary transmission among adults, especially women continuous to decline after recovery especially after proper administration of HAART (*σ*_1_ = 0.25, *σ*_1_ = 0.85, *ρ*_1_ = 0.012, *ρ*_2_ = 0.35) and this will also affect the recovery rate in the juvenile sub-population because there will be less secondary transmission from Mother-To-Child. However, when the probability of secondary transmission among adult sub-population is high, the recovery rate continues to increase over a relatively long period of time and declines after a certain period despite the administration of HAART treatment as can be seen in Figure 4.3 because as much as efforts is made to control the spread of HIV/AIDS in children who have no incidence rate (*B*_1_(*t*) = 0) there is need to control the secondary transmission rate of the disease in adults before treatment is embarked upon. However, failure to control the secondary transmission among children will render the efforts to curb the rise in prevalence among children useless. Also, recovered juvenile sub-population would be exposed to the disease further instead of moving to the undetectable=untransmitable compartment.

**Figure 4.1.**
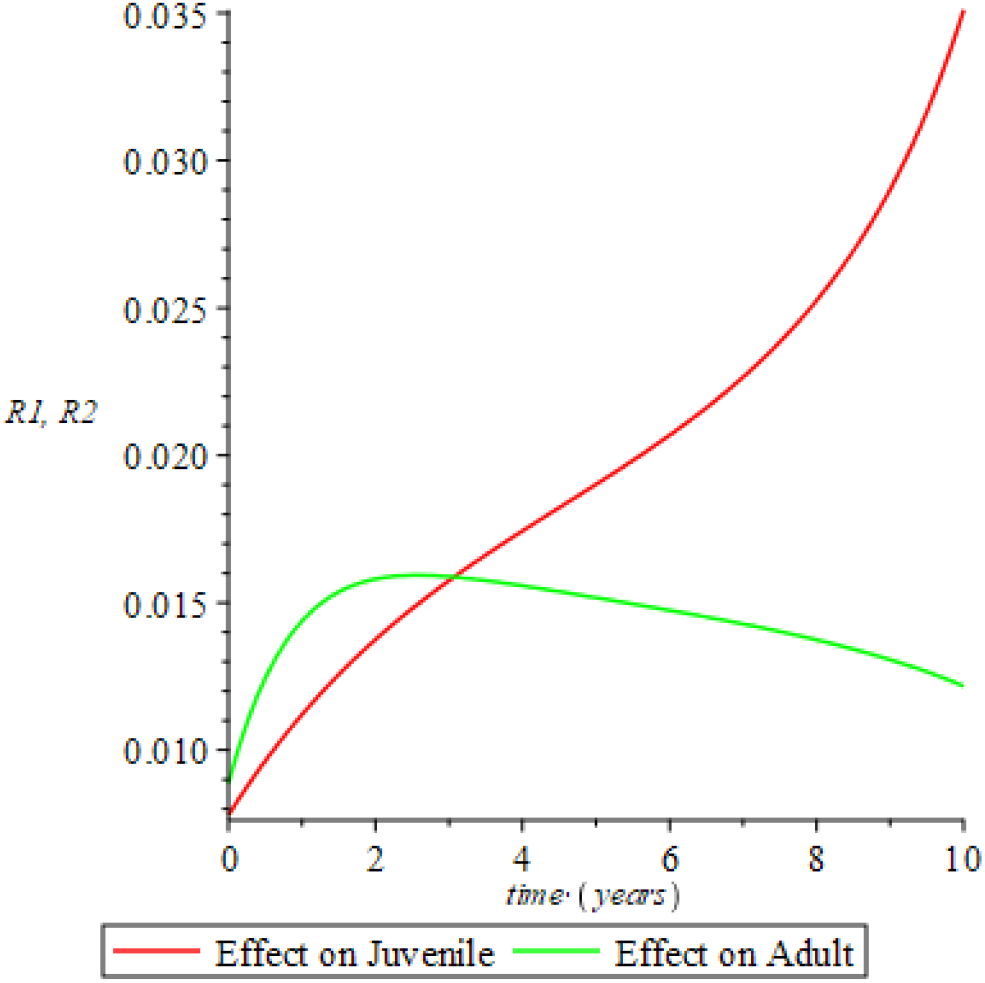
Recovery in the Juvenile sub-population when the probability ofsecondary transmission is low in the adult sub-population (*σ*_1_ = 0.25, *σ*_1_ = 0.85, *ρ*_1_ = 0.012, *ρ*_2_ = 0.35).

**Figure 4.2.**
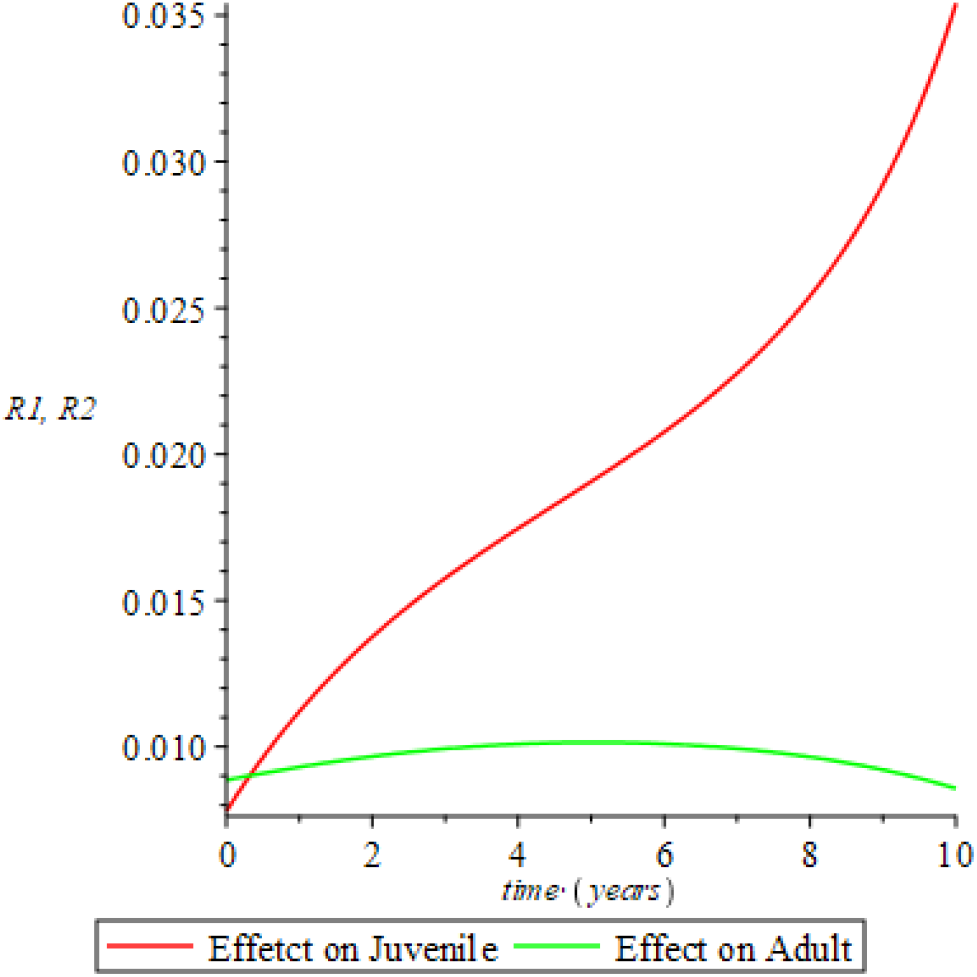
Recovery in the Adult sub-population when the probability of transmission is low in the adult sub-population (*σ*_2_ = 0.25, *σ*_2_ = 0.75, *ρ*_1_ = 0.012, *ρ*_2_ = 0.35)

**Figure 4.3.**
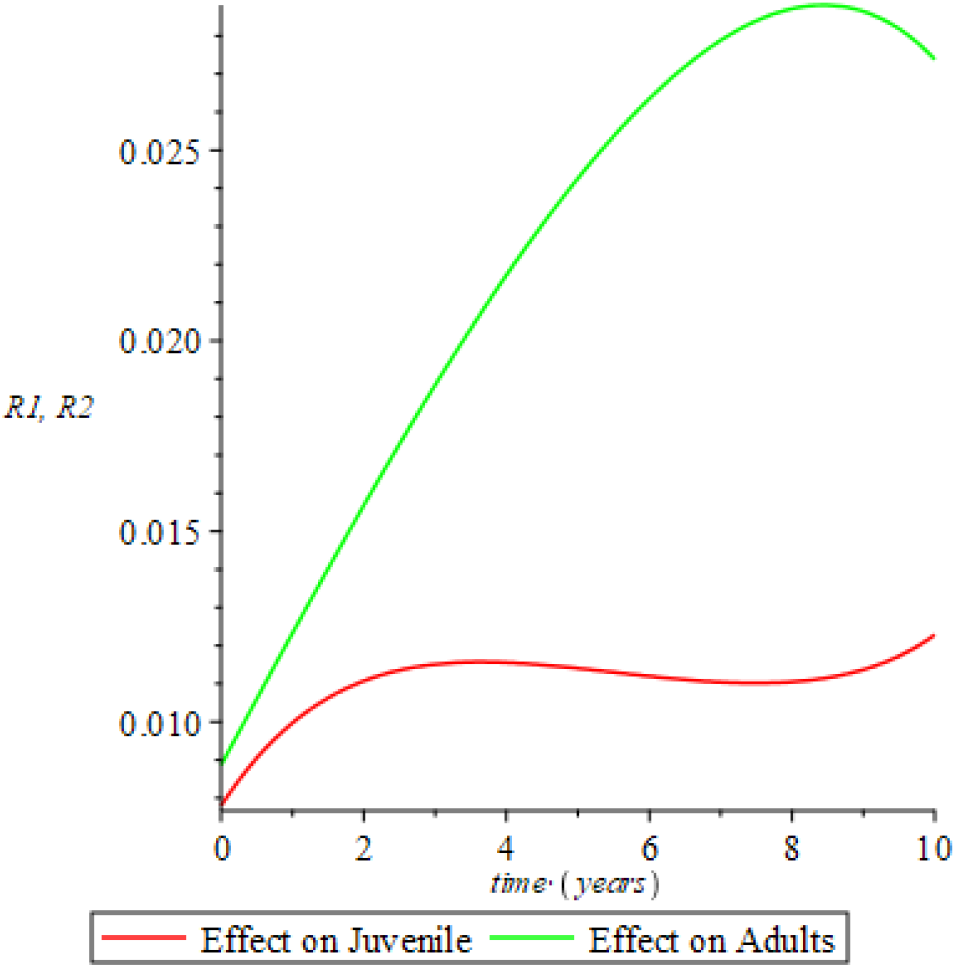
Recovery in the Juvenile sub-population when the probability of transmission is high in the adult sub-population (*σ*_1_ = 0.25, *σ*_1_ = 0.85, *ρ*_1_ = 0.150, *ρ*_2_ = 0.010)

Meanwhile, in Experiment 2 the effect of treatment on the recovery in the adult sub-population was investigated as shown in Figure 4.2 when the probability of transmission is low. Result shows clearly that the recovery rate rises progressively over a long period among the adult sub-population when the probability of secondary transmission is low from the recovered compartment actively administering HAART consistently over a long period of time as seen in Figure 4.2. Therefore, to enhance the movement of adult sub-population in an endemic situation from the recovered compartment to the undetectable compartment over a long period of time, the probability of transmission must be reduced to the barest minimum or possibly negligible. This can be achieved by adequately administering the HAART treatment as well creating awareness among adult sub-population on safe sex behaviors and other behavioral changes as required to reduce the risk of contracting or re-contacting the disease. In other to achieve the goal of [3], the rate of secondary transmission after recovery by the recovered compartment actively administering HAART must be zero and hence according to this study, it is found that the Reproductive number is zero (*R*_0_ = 0) since there is a zero incidence rate among the juvenile sub-population.

Comparing Figure 4.4 and Figure 4.5 with Figure 4.6 and Figure 4.7 while investigating the effect of treatment when the secondary transmission rate is low and high respectively. As shown in Figure 4.4 and Figure 4.5 when the probability of secondary transmission is low and treatment rate is high, then the proportion of recovery in the adult sub-population is drastically reduce after the 3^rd^ year and the re-infection rate persists in small proportion, while the re-infection rate in the juvenile sub-population is eradicated. Therefore, when the probability of secondary transmission is high and treatment rate is high then recovery rate will increase accordingly as seen in Figure 4.6 and 4.7 respectively. This clearly shows the effect of uncontrolled sexual behaviours among adults sub-population which makes their incidence rate *B*_2_(*t*) > 0 unlike in the case of the juvenile sub-population with no incidence rate or chances of children to children transmission. As a result, despite the high treatment rate among adult sub-population and low probability of transmission, the behavioral change is important to completely eradicate the disease among adults.

**Figure 4.4.**
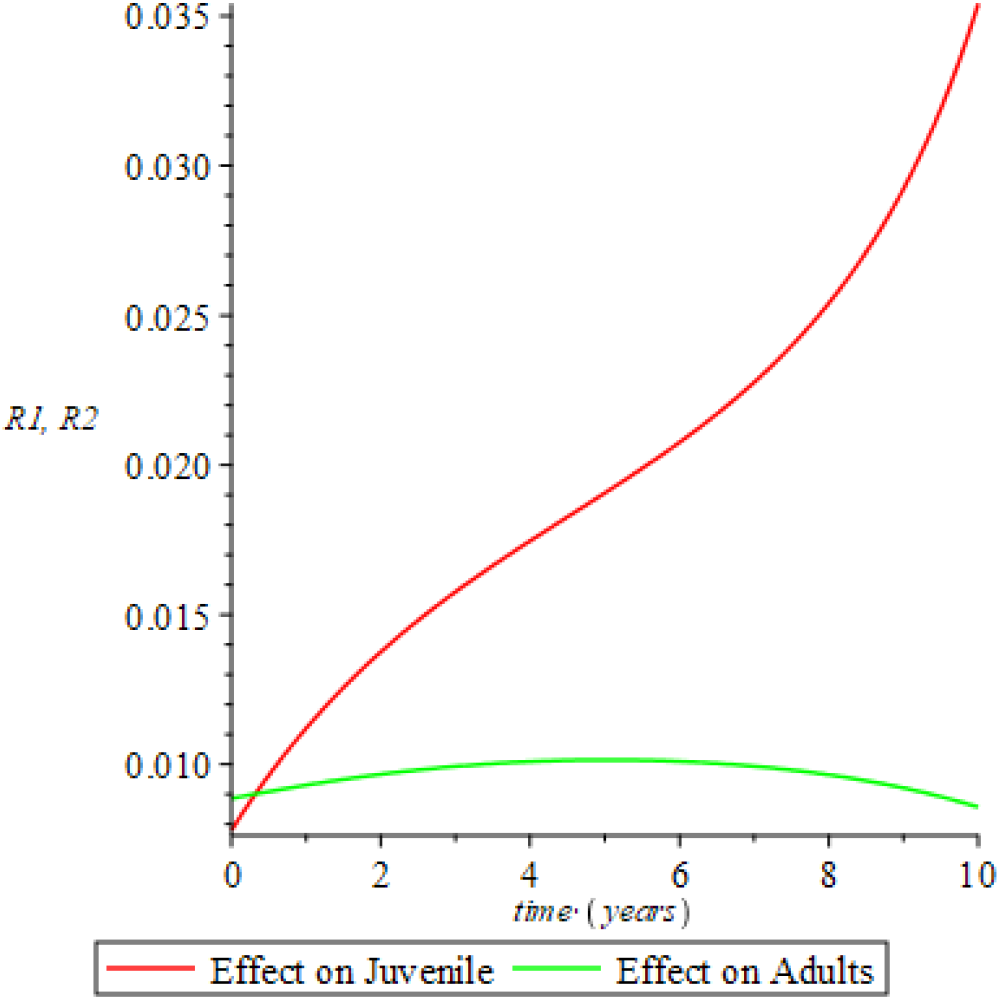
Recovery when the treatment rate is low and the probability of transmission is low (*σ*_1_ = 0.25, *σ*_2_ = 0.25, *ρ*_1_ = 0.012, *ρ*_2_ = 0.35)

**Figure 4.5.**
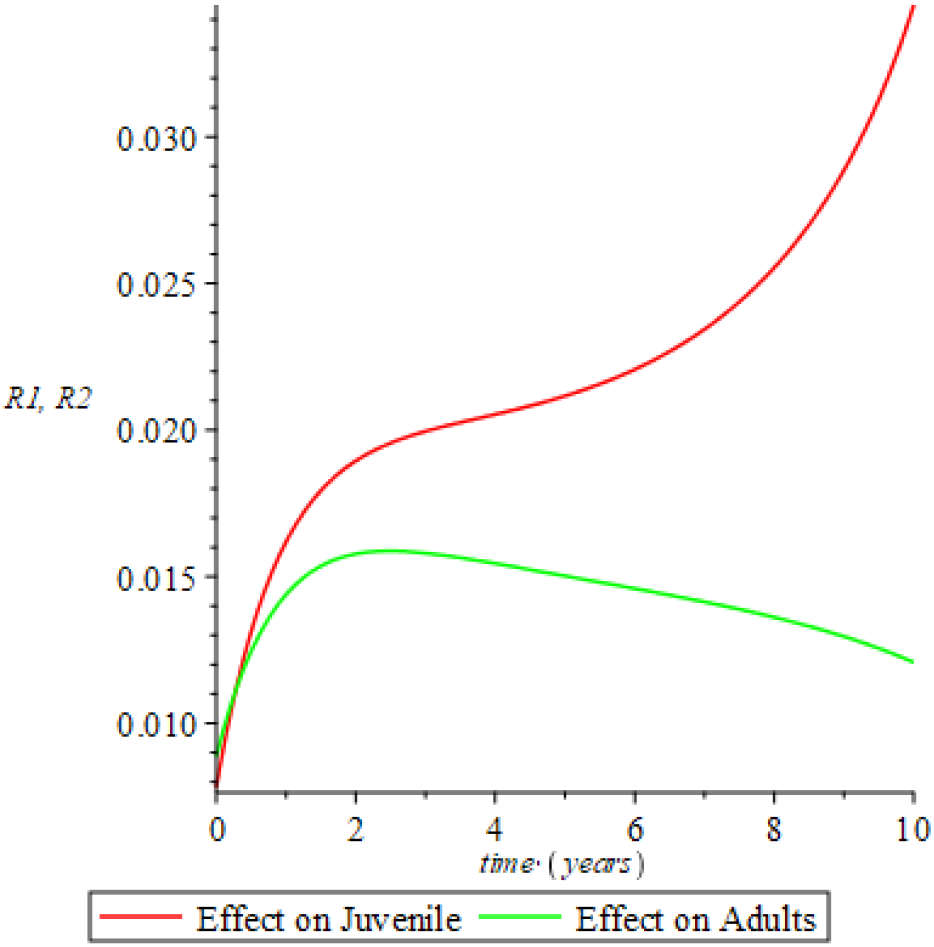
Recovery when the treatment rate is high and the probability of transmission is low (*σ*_1_ = 0.85, *σ*_2_ = 0.85, *ρ*_1_ = 0.012, *ρ*_2_ = 0.35).

**Figure 4.6.**
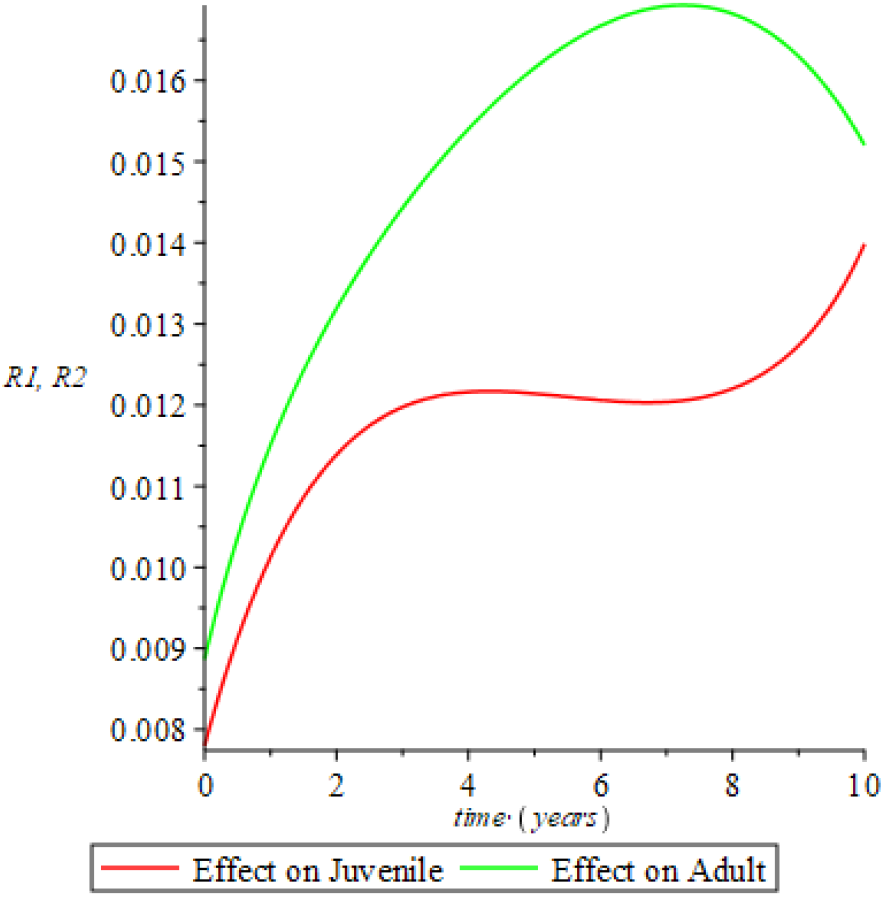
Recovery when the treatment rate is low and the probability of transmission is high and (*σ*_1_ = 0.25, *σ*_2_ = 0.25, *ρ*_1_ = 0.150, *ρ*_2_ = 0.010).

**Figure 4.7.**
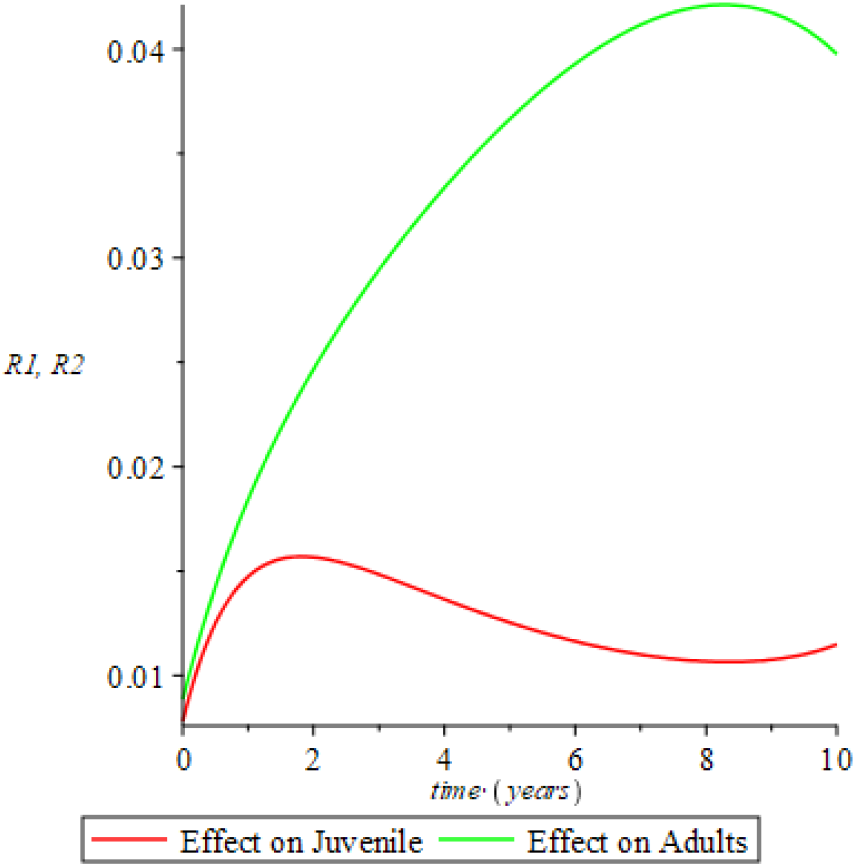
Recovery when the treatment rate is high and probability of secondary transmission is high (*σ*_1_ = 0.85, *σ*_2_ = 0.85, *ρ*_1_ = 0.150, *ρ*_2_ = 0.010)

The effect of treatment on recovery of adult sub-population when the juvenile sub-population is left untreated is presented as Experiment 8 and 9 shows that the treatment of adult sub-population enhances recovery in both adult and juvenile sub-population even specifically when treatment rate is high and transmission rate is low. In Figure 4.8, when the treatment rate in the adult sub-population is high and the juvenile sub-population is left untreated, there is stiff recovery among the adult sub-population over a short period of time before it begins to decline due to the maturation of the juvenile sub-population who are left untreated. As such when the untreated juvenile sub-population matures to adult sub-population, their incidence rate becomes greater than zero and hence re-infection occurs among the adult sub-population which in turn slows down recovery. And in Figure 4.9, it is evident that as treatment rate increase in the adult sub-population there is sharp rise of recovery in a long run. Hence, it is significant that both juvenile and adult sub-population undergo active vaccination and preventive procedures at the same time to ensure the full control of cases of secondary infection from the recovered compartment and as a result, the possible eradication of the disease even in an endemic situation. In Experiment 10 when the rate of treatment is low and the probability of secondary transmission after recovery is low and the adult sub-population is left untreated, the recovery rate in the adult sub-population experiences a sharp decline in a short run as shown in Figure 4.10 and increase in adult sub-population since there appears to be zero rate of incidence among children to children. Meanwhile, as in Experiment 11, when the treatment rate is high and the probability of secondary transmission is high when the adult sub-population is left untreated the adult recovery rate declines swiftly and it therefore means the secondary infection rate of the adult sub-population also increases rapidly due to the high incidence rate (see Figure 4.11) where as high treatment rate enhance high recovery among the juvenile sub-population.

**Figure 4.8.**
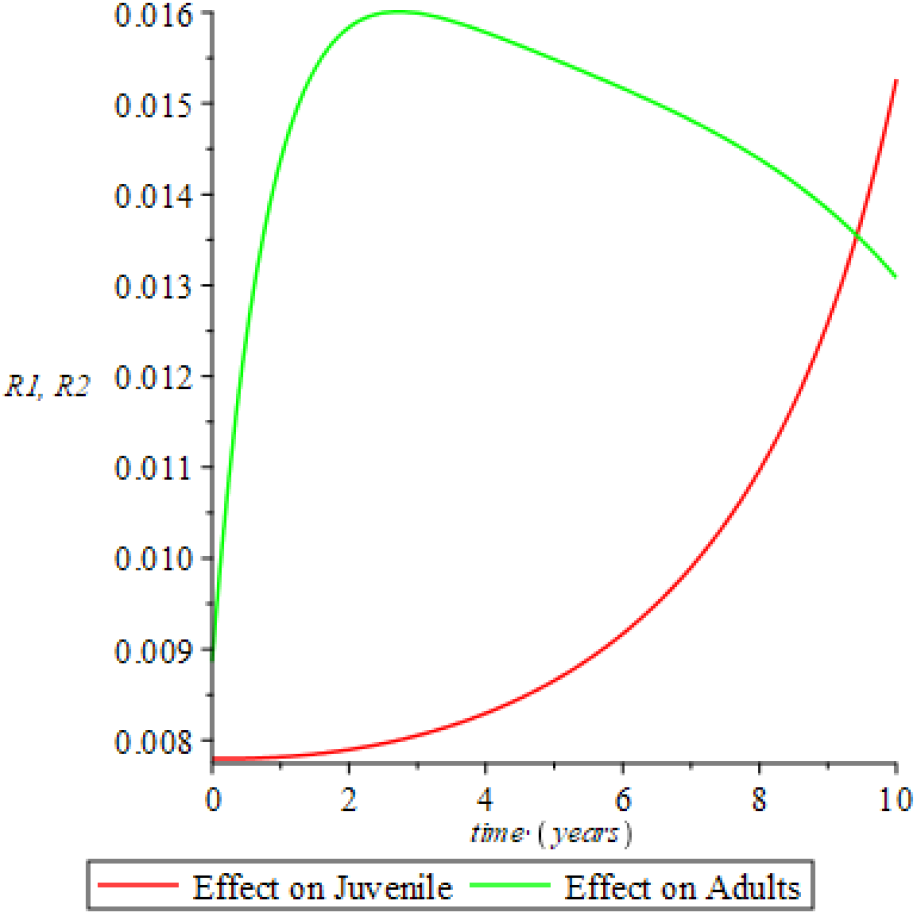
Recovery with high treatment rate when the probability of transmission is low and the juvenile sub-population is left untreated (*σ*_1_ = 0.00, *σ*_2_ = 0.85, *ρ*_1_ = 0.012, *ρ*_2_ = 0.35).

**Figure 4.9.**
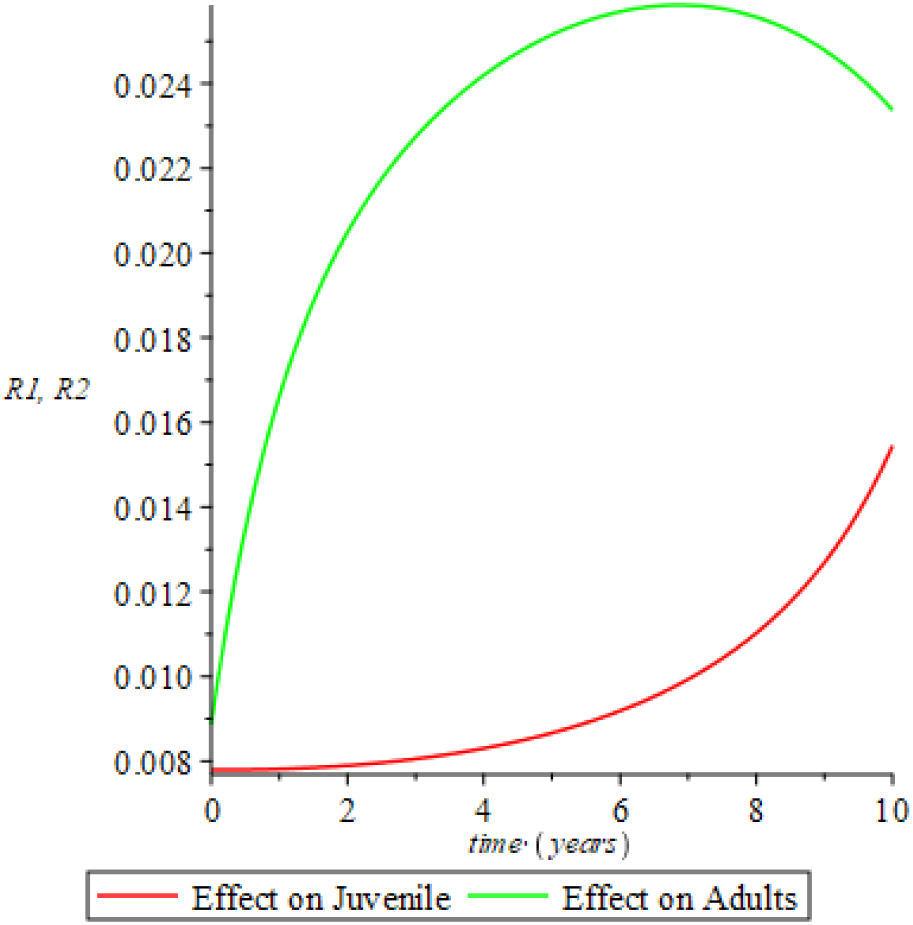
Recovery with high treatment rate when the probability of transmission is high and the juvenile sub-population is left untreated (*σ*_1_ = 0.00, *σ*_2_ = 0.85, *β* = 0.150, *β*^′^ = 0.010).

**Figure 4.10.**
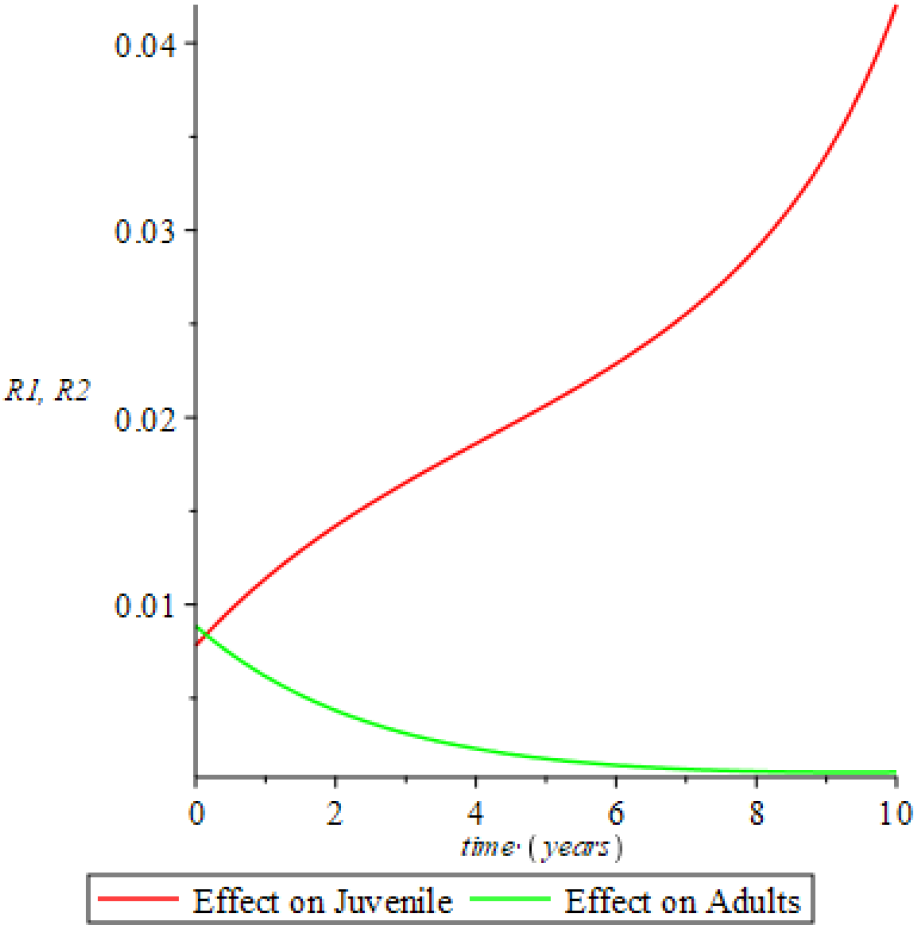
Recovery with low treatment rate when the probability of transmission is low and the adult sub-population is left untreated (*σ*_1_ = 0.25, *σ*_2_ = 0.00, *ρ*_1_ = 0.012, *ρ*_2_ = 0.35)

**Figure 4.11.**
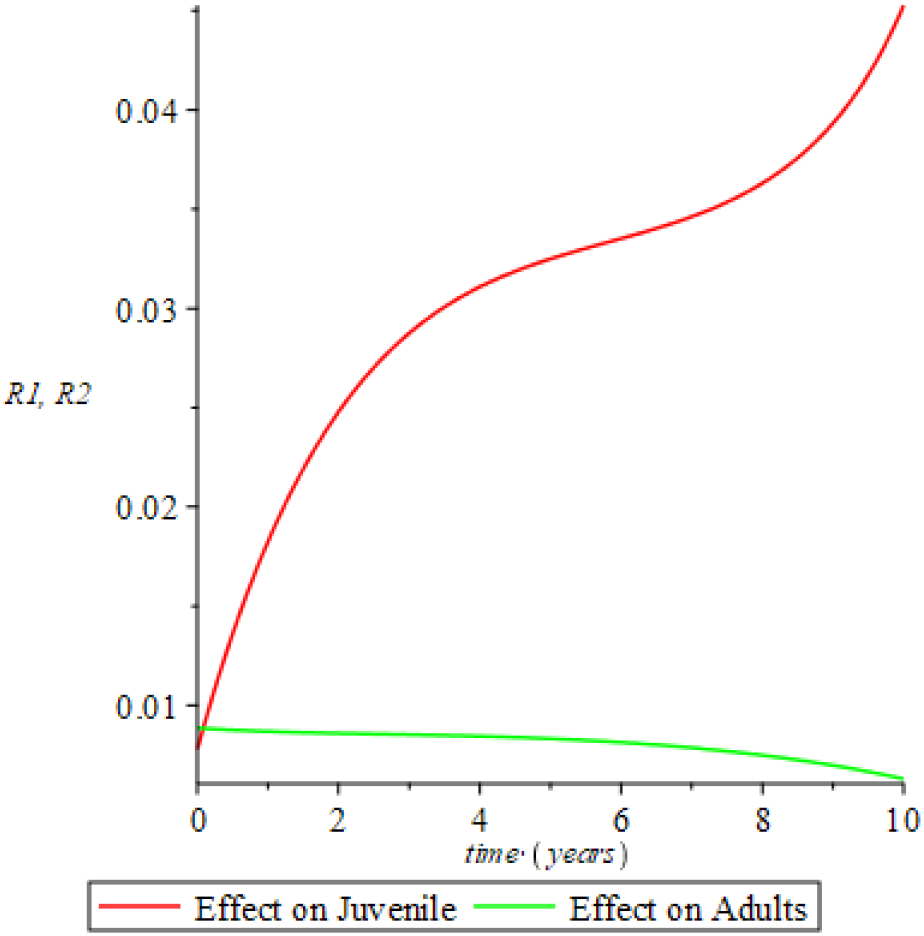
Recovery with high treatment rate when the probability of transmission is high and the adult sub-population is left untreated (*σ*_1_ = 0.85, *σ*_2_ = 0.00, *ρ*_1_ = 0.150, *ρ*_2_ = 0.010).

However, the effect of vertical transmission (also referred to as Mother-to-Child transmission) was also captured in Experiment 12, 13 and 14. When all the newborns from infected mothers are all HIV positive (*ξ* = 1) as shown in Figure 4.13 and Figure 4.13, the recovery rate drops slowly among juvenile but increases among adult sub-population when treatment rate is both low and high respectively for both age-structures.

In conclusion, in other to understand the impact of different proportions of recovered juveniles moving to the undetectable compartment in the juvenile sub-population when treatment rate is low and high in Nigeria, Experiment 15 in Figure 4.15 illustrates that high treatment rate is necessary to control vertical secondary transmission of the disease, especially when the probability of transmission is negligible.

**Figure 4.12.**
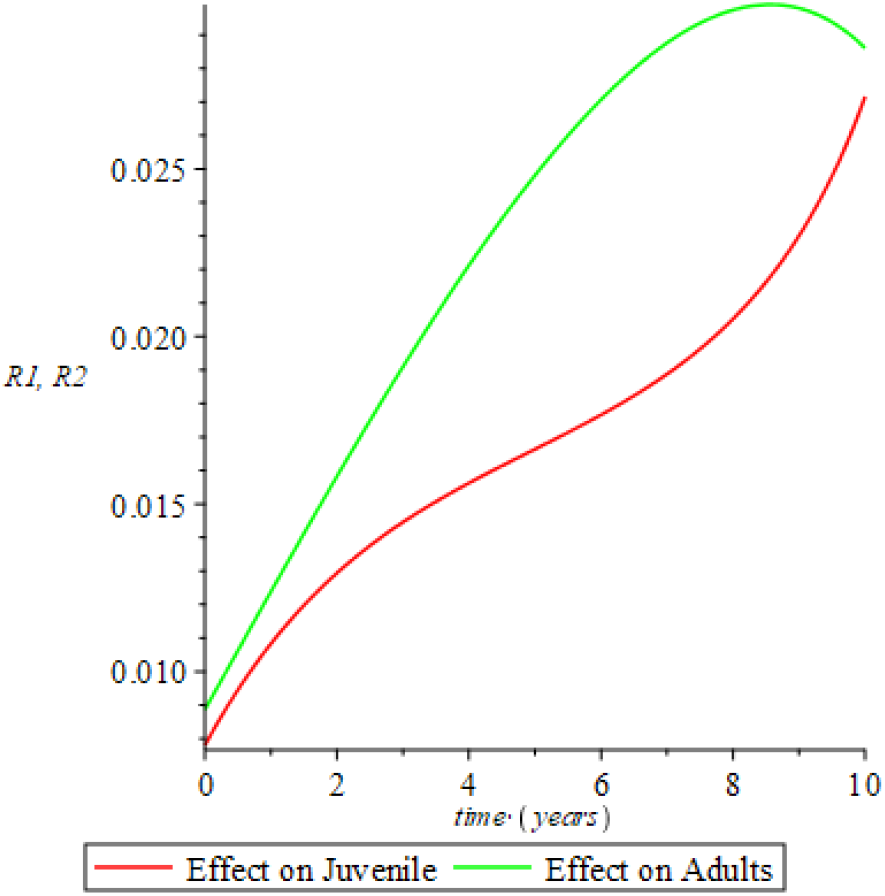
Recovery in the juvenile and adult sub-population when all newborns from infected mothers are HIV positive and treatment rate is low. (*σ*_1_ = 0.25, *σ*_2_ = 0.25, *ρ*_1_ = 0.075, *ρ*_2_ = 0.005, *ξ* = 1.0).

**Figure 4.14.**
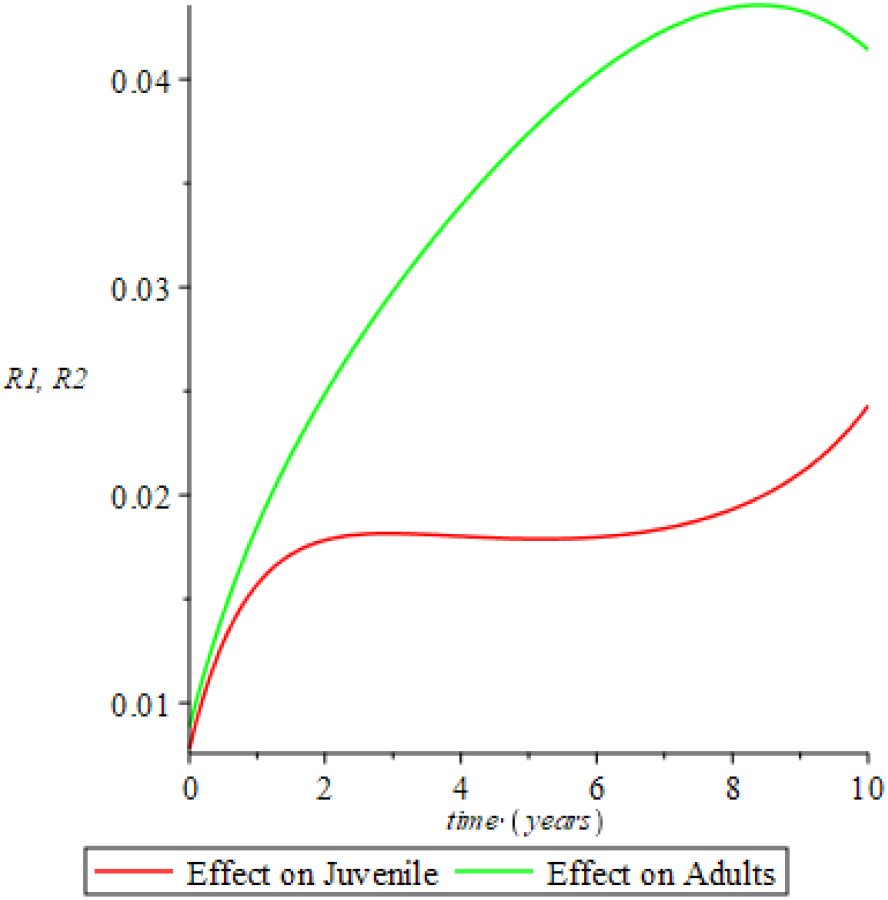
Recovery in the juvenile and adult sub-population when all newborns from infected mothers are HIV positive and treatment rate is high (*σ*_1_ = 0.85, *σ*_2_ = 0.85, *ρ*_1_ = 0.075, *ρ*_2_ = 0.005, *ξ* = 1.0).

**Figure 4.15.**
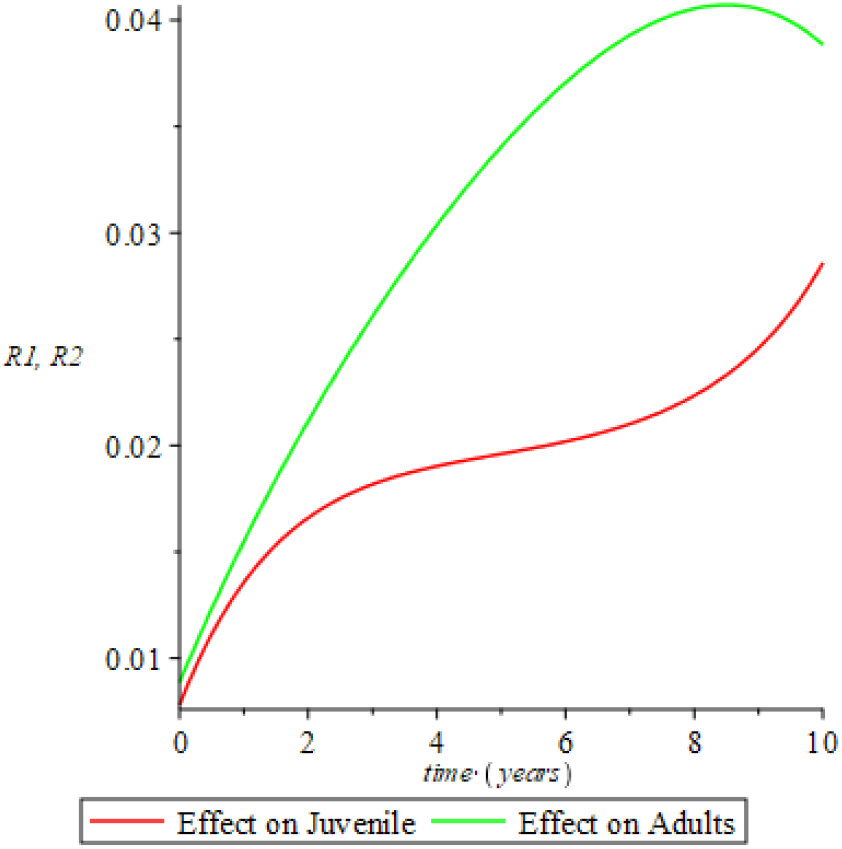
Recovery in the juvenile sub-population showing the effect of vertical transmission when all newborns from infected mothers are HIV negative and HIV positive respectively. (*σ*_1_ = 0.5, *σ*_2_ = 0.5, *ρ*_1_ = 0.075, *ρ*_2_ = 0.005, *ξ*_+_ = 1.0, *ξ*_−_ = 0.0).

Here, models that captures preventive strategies in an endemic situation based on stability analysis which prevent further incidence rate of the disease and consequently provides model conclusion for the eradication of the disease if the population in the undetectable=untransmitable compartment continue actively with the HAART treatment and have a controlled sexual behaviors is formulated and implemented for trial control in other to enhance strategies for HI/AIDS eradication in Africa.

## 5. CONCLUSIONS

In this paper results from the implementation of the model data of Nigeria shows that there is a need to enhance the availability and active use of the HAART treatments to both infected and recovered juvenile and adult members of the *I*-class and *R*-class respectively as this would not only increase the prevalence of infection but also the recovery rate if risky sexual behaviours among heterosexual adults is not controlled. Behavioral interventions is key based on findings from this research as well as active administration of the HAART treatment in other to control secondary re-infection in an endemic situation where vertical transmission is in control and the assumption of zero immigration is true.

If the model threshold parameters are satisfied without loose of generality, the spread and secondary transmission of HIV/AIDS among heterosexual adults and Mother-to-Child transmission can be controlled and possibly eradicated as this study suggests with the secondary Reproductive Number being zero (*R*_0_ = 0) there would not be further infection in the adult sub-population and the juvenile sub-population incidence rate is also zero (*B*_1_(*t*) = 0). Also, as much as efforts are on the way to attain U=U in Nigeria, the active administration of HAART treatment on juvenile and adults sub-population is very key and when sufficiently and actively administered would ensure that the transmission rate between infective and recovered individuals is zero. An eradication of secondary transmission in a close population to attain a zero reproductive number can be achieved through active administration of HAART, high and target health education, counselling and testing as well as behavioral change of all the classes of the SEIRUS compartment.

Also, as much as the rate of vertical transmission has been considerably reduce to about 5% in Africa, the role of Mother to Child (MTC) transmission cannot be ignored because the rate of maturation from juvenile to adult in the susceptible class *η*_*s*_, exposed class *η*_*e*_, infectious class *η*_*i*_, and the recovered class *η*_*r*_, affects the endemic equilibrium state of the entire population. With the new Reproductive Number *R*_0_ = 0 then the probability of transmission from adult to adult and mother to child is reduced to zero and the susceptible juvenile must retain their HIV negative status as they mature to adults, also the recovered juvenile must retain their HIV negative status as they mature to adult in other to move into the undetectable class of the adult sub-population. The maturation rate of susceptible and recovered juvenile must be taken into consideration when planning strategies for control and eradication of the HIV/AIDS disease in Africa as well as retaining the HIV-negative status of both the juvenile and adult sub-populations.

## Data Availability

The data available for use in this study was retrieved from WPR (2019) and UNAID (2017)

## REFERENCES

[1] Oduwole, H. K. and Kimbir, A. R. (2018). “Modelling vertical transmission and the effect of Antiretroviral Therapy (ART) on the dynamics of HIV/AIDS in an age-structured population in Nigeria. Journal of Natural and Applied Science-Nasara Scientifique, Vol. 7 no. 1 pp. 51–78.

[2] Mugisha, J.Y.T. and Luboobi, L.S. (2003). Modelling the effect of vertical transmission in the dynamics of HIV/AIDS in an age-structured population. S. Pac. J. Nat. Sci., 21 (B), 82–90

[3] Centers for Disease Control and Prevention (CDC). (2018, 31, August). Comprehensive Prevention Programs for Health Departments. www.cdc.gov/hiv/programresources/healthdepartments.

[4] Contag, C.H., Ehrnst, A., Duda, J., Bohlin, A.B., Lindgren, S., Learn, G.H., and Mullins, J.I. (1997). Mother-to-infact transmission of human immunodeficiency virus type 1 involving five envelop sequence subtypes. J. Virol. 71:1292–1300

[5] Diekmann, O, Heesterbeek, J. A. P and Roberts, M. G. (2009); The construction of next-generation matrices for compartmental epidemic models, J. R. Soc. Interface, 2009 (doi:10.1098/rsif.2009.0386)

[6] Van den Driessche, P. and Watmough, J. (2002); Reproduction numbers and subthreshold endemic equilibria for compartmental models of disease transmission. Math. Biosci. 180, 29–48, 2002 doi:10.1016/S0025-5564(02)00108-6

[7] World Population Prospects (2019 Revision) - United Nations population estimates and projections. Total population: Estimated to be consistent with the 1963, 1991 and 2006 censuses, adjusted for underenumeration, with the age and sex structure from the 2011 MICS4 survey, and with estimates of the subsequent the trends in fertility, mortality and international migration.

[8] World Health Organization (WHO) (2018). Life expectancy in South Africa. [pdf]

[9] UNAIDS (2017). A snapshot of men and HIV in South Africa.[pdf]

[10] UNAIDS (2012). “The quest for an HIV vaccine” http://www.unaids.org/en/resources/presscentre/featurestories/2012/may/20120518vaccinesday/

[11] Kgosimore, M. and Lungu, E.W (2006) The effects of vertical transmission the spread of HIV/AIDS in the presence of treatment. Mathematical Biosciences and Engineering, 3, 297–312. doi:10.3934/mbe.2006.3.297

